# Multi-criterion uncertainty estimation improves skin cancer distribution shift detection and malignancy prediction

**DOI:** 10.1101/2025.08.20.25334101

**Authors:** W. Max Schreyer, Ravi Samatham, Elizabeth Berry, Reid F. Thompson

## Abstract

Widespread access to imaging technologies and stronger machine learning (ML) architectures for dermatology tasks such as malignancy prediction have spurred a race to develop models to assist in the automated diagnosis of skin cancer. However, high diagnostic performance on benchmarking datasets quickly deteriorates when models are challenged with data from disparate clinical sources. Generalization gaps stem from the high variability in skin lesion images due to lighting, capture angle, imaging technology and patient phenotype among other factors, impeding the safe application of diagnostic ML models in practice. In this study, we apply a novel multi-criterion uncertainty-estimation approach to detect out-of-distribution skin lesion images from four publicly available datasets across five countries. Using our method, Supervised Autoencoders for Generalization Estimates (SAGE), we quantify likeness of images from patients in Argentina, Brazil, Austria, North Macedonia, Turkey, Australia and the United States to the popular HAM10000 benchmarking dataset and identify problematic image artifacts affecting the reliability of predictions in a pre-clinical setting. We show how filtering images based on SAGE score thresholds can improve the performance of a separate malignancy prediction model and how our approach is robust to variations in image modality and the introduction of new diagnostic classes, providing users with a powerful tool for interrogating key differences between their data and the training distribution of an ML model before clinical implementation.

## Introduction

Skin cancers are the most commonly-diagnosed malignancy worldwide, with over 300,000 cases of melanoma (age-standardized incidence rate (ASIR) = 3.56 per 100,000 people), 1.9 million cases of squamous cell carcinoma (ASIR = 22.38) and 4.4 million cases of basal cell carcinoma (ASIR = 74.10) occurring globally in 2021.^1,2^ However, malignancies represent only a fraction (22%) of all index skin lesions referred to dermatologists for biopsy and review, resulting in a clinically significant tension between diagnostic sensitivity and specificity.^3^ Diagnostic capabilities may also vary among primary care providers (PCPs) and other skin-screening clinicians according to level of training, years of experience, and use of dermoscopy.^4^ Further barriers to skin cancer detection combine to exacerbate pre-existing health disparities such as patients with low socioeconomic status also living in rural locations.^2,5^ Fortunately, the combination of widely available imaging technologies and automated detection through machine learning (ML) have the potential to revolutionize skin cancer diagnosis and improve global health equity as a result.^6,7^

Many ML models have been trained to identify cutaneous malignancies within skin imaging datasets.^8–16^ However, there are notable differences between highly-curated benchmarking datasets used to train and evaluate ML models and the images encountered in real world clinical settings, often leading to reduced model effectiveness in practice.^4,17,18^ Even state-of-the-art models suffer from these performance drops; the top 25 entries from the ISIC 2019 Grand Challenge misclassified nearly 50% of previously-unseen skin lesion images despite stellar performance on benchmarking datasets.^19^ Ambitious ML-assisted dermatology smartphone phone apps have also been shown to have low accuracy and can therefore pose a harm to users attempting to self-diagnose potentially life-threatening diseases without expert oversight.^20–22^ One major source of difficulty in automating skin cancer diagnosis is the variability in imaging technologies like dermatoscopes which cannot properly standardize inputs to predictive algorithms – outputs can vary based on whether or not the dermatoscope is polarized and the type of light source.^23,24^ Indeed, even the presence of modest artifacts such as blur or blue/red shifted-pixel intensities showed marked decreases to ML performance for both diagnostic and disease management tasks when compared to control images.^25^ Beyond device or light-induced inconsistencies, patient phenotypes such as dark skin have been shown to degrade ML model accuracy and pose potential ethical dilemmas in the deployment of algorithms trained on benchmarked datasets which either omit skin type metadata or severely underrepresent some patient populations.^26,27^ It is critically important to assess the similarity of new dermatological images with a model’s training data on a case-by-case basis, in order to reduce disparities and identify potentially problematic samples before a model is used in any treatment pipeline.

Some diagnostic dermatology models have attempted to remove corrupted or low-quality samples through out-of-distribution (OOD) detection or by building inherent uncertainty quantification (UQ) into the prediction task.^28–30^ This typically combines UQ with another primary task, lacking flexibility in adapting to different tasks or pairing with stronger models as they become available. Other single-pronged approaches to measuring uncertainty such as using maximum classifier softmax outputs can be intrinsically measured at time of prediction, but fail silently under scenarios of data drift.^31^ Furthermore, distribution shift occurs across a spectrum where semantically unrelated examples, new classes (e.g. different diagnoses), altered class proportions and image corruptions may increase the difficulty of OOD detection and selective prediction tasks.^32^

Our previous work has introduced an approach (Supervised Autoencoder for Generalization Estimates, SAGE) which derives a composite UQ score from both supervised and unsupervised measures of epistemic uncertainty.^33^ SAGE therefore acts as both an OOD detector when test examples are scored and compared to training data, and a decoupled metric for selective prediction which can prevent examples with a high risk of failure from being evaluated by a downstream model. In this study we implement SAGE to assess the generalization potential of a skin cancer detection algorithm^26^ across diverse dermatology images including OOD examples. We show that our SAGE scoring system improves OOD detection performance over several widely used methods and enables selective prediction for in-distribution (ID) images, enriching for examples that are appropriate versus problematic for ML-assisted diagnosis.

## Methods

### Datasets

**Humans Against Machine (HAM10000)**^34^ consists of 10,015 dermoscopy photos sourced from the Rosendahl dermatological practice in Australia and the ViDIR Group in Austria. We downloaded files from the Harvard Dataverse (https://doi.org/10.7910/DVN/DBW86T) containing JPEG images and associated metadata as a CSV file. The metadata file contains columns detailing lesion and image identifiers, patient age at time of image capture (median = 50 years old), body site of the lesion and diagnostic method. All images are of pigmented lesions and ground-truth labels are included in the metadata with six diagnostic classes: actinic keratosis (*ak*), basal cell carcinoma (*bcc*), benign keratosis (*bk*), melanocytic nevus (*nevi*), melanoma (*mel*) and vascular skin lesion (*vasc*). For the purposes of this study, only *bcc* and *mel* classes were considered malignant; all other classes were considered benign, although *ak* is known to progress to cancer in certain cases. All malignant lesions were histologically-confirmed. Images have an original size of 600x450 pixels and were resized using bilinear interpolation where the smaller dimension of height or width was resized to 299 pixels, preserving aspect ratio. Images were cropped to 299x299 pixels after resizing to ensure consistency across datasets and compatibility with open-source ML architectures. Authors include multiple images of the same lesion with differing perspectives as a form of natural data augmentation and perform quality control to remove out-of-focus images and images with insufficient zoom of small lesions. We randomly-split HAM10000 into training and test sets stratified by diagnostic class and removed any images of lesions from the test set that were also present in the training set, leaving 9,013 images for training and 600 for testing.

The **Hospital Italiano de Buenos Aires (HIBA)**^35^ dataset contains 1,616 JPEG images of mixed dermoscopic and clinical smartphone images from Argentina which were downloaded from the ISIC archive (https://doi.org/10.34970/587329). The metadata file contains a row for each image which includes a unique image and lesion ID as well as an anonymized patient identifier. Separate metadata columns are also included for age (median = 65 years old), sex, family history of skin cancer, Fitzpatrick Skin Type (FST)^36^ and image type differentiating between clinical smartphone and dermoscopic imaging technologies. 93% (*n*=566) of the 623 patients had skin type data and age and sex were recorded for over 99% of patients. Each image is assigned a ground-truth label from one of 10 diagnostic classes, all of which overlap with HAM10000 diagnostic classes. We sort HIBA images into 8 harmonized image classes: basal cell carcinoma (*bcc*), melanoma (*mel*), squamous cell carcinoma (*scc*), melanocytic nevus (*nevi*), actinic keratosis (*ak*), dermatofibroma (*df*), vascular skin lesion (*vasc*), solar lentigo (*bk*), seborrheic keratosis (*bk*) and lichen planus-like keratosis (*bk*). All malignancies for *bcc, scc* and *mel* lesions were biopsy-confirmed. Images from the HIBA dataset vary in size between a maximum of 4128x3096 pixels and a minimum of 162x152 pixels. All files were preprocessed by converting to 8-bit RGB color channels, resized with bilinear interpolation and center-cropped to 299x299 pixels before use in our study.

The dermatology program at the **Universidade Federal do Espírito Santo (UFES)**^37^ in Brazil published a dataset in 2020 containing 2,298 clinical smartphone images with 6 diagnostic classes: basal cell carcinoma (*bcc*), squamous cell carcinoma (*scc*), actinic keratosis (*ak*), melanoma (*mel*), melanocytic nevus (*nevi*) and seborrheic keratosis (*bk*). All *bcc, scc* and *mel* images were considered malignant and lesions were biopsy-confirmed. We downloaded images in PNG format and the metadata CSV file from the Mendeley Data Commons link provided by the authors (https://doi.org/10.17632/zr7vgbcyr2.1). The metadata file contains patient, lesion and image identifiers as well as detailed lifestyle and living condition information such as access to piped water and sewage systems, smoking, drinking and exposure to pesticides. 100% of images have a skin phototype using FST while only 65% of images contain patient sex. The median patient age is 62 years old with a minimum age of 6. During manual review of UFES data, we found 43 images with inconsistent or mixed labeling (e.g. duplicate images assigned to different ground-truth diagnoses: 6 instances where the ground-truth label switches between *scc* and *bcc* and 7 other instances where the label change impacts designation of the lesion as benign/malignant. All mislabeled examples were removed prior to analysis. Images from the UFES dataset have varying sizes with a maximum of 3474×3476 pixels and a minimum of 147×147 pixels. All files were converted to 8-bit RGB color channels, resized using bilinear interpolation and center-cropped to 299×299 pixels before use.

The **Diverse Dermatology Images (DDI)**^26^ dataset consists of 656 images from Stanford dermatology clinics in the US captured between 2010 and 2020. All photos were taken on a clinic-issued smartphone and extracted retrospectively from electronic health records. We downloaded images and the associated metadata file from the Stanford AIMI Datasets Azure link (https://stanfordaimi.azurewebsites.net/datasets/35866158-8196-48d8-87bf-50dca81df965). The DDI dataset features over 40 unique diagnoses, including examples from all HAM10000 classes, but 36 others with low incidence in the general population and not contained in the other imaging datasets included in our analysis. (Extended Table 2) Metadata for DDI images is sparse with only a unique image identifier, diagnosis, malignancy and skin type information included for each lesion. Notably, sex and unique patient or lesion identifiers were absent. Images had varying sizes with a maximum of 1914×1424 pixels and a minimum of 163×79 pixels. We preprocessed DDI by converting images to 8-bit RGB color channels, resizing using bilinear interpolation and center-cropping to 299×299 pixels before use. Authors of DDI performed additional data augmentation including random rotation and vertical flipping of images before training their malignancy prediction model, however we were unable to exactly replicate this process due to lack of access to the model’s training code.

The **MILK10K** ^38^ dataset is part of the ongoing 2025 ISIC benchmarking challenge and consists of 10,480 images from 5,240 dermatology cases sourced from Australia, Austria, North Macedonia, Turkey and the United States. Each MILK10K case contributes one dermoscopic and one clinical smartphone image and detailed metadata is provided by the authors containing patient approximate age, anatomic site of the lesion, automatically extracted skin tone and 11 simplified and hierarchical diagnostic classes. We downloaded benchmark images in JPEG format and associated metadata from the provided Harvard Dataverse link (https://dataverse.harvard.edu/dataset.xhtml?persistentId=doi:10.7910/DVN/FSXRAQ). 95.7% (n = 5016) of the 5,240 cases have a biopsy-confirmed diagnosis. All images are 600×450 pixels in size and we apply a crop to 299×299 pixels for use.

**Caltech101**^39^ is a classical machine learning dataset with 9,144 RGB color image examples from 101 object categories (e.g. “accordion”, “airplane”, “anchor”…), which we use for the purpose of far-OOD detection with non-semantic data shift. Images were originally sourced from Google Images and resolution is variable, although most examples are 300×200 pixels. We downloaded the dataset from Caltech Data (https://data.caltech.edu/records/mzrjq-6wc02) and apply center cropping (299×299 pixels) with pixel normalization before use in UQ and malignancy prediction pipelines.

### Model Training

We first normalized all input pixel values using mean and standard deviations from the ImageNet dataset. All image preprocessing was completed using pillow (v.10.3.0) and the torchvision (v0.17.2) transforms library. The SAGE model architecture was written in python (v3.10.14) using pytorch^40^ (v2.2.2) and consists of an encoder as well as decoder and classifier modules that take the encoder’s compressed embedding as input. The decoder contains 2 fully-connected layers, 6 deconvolutional layers and a learned upsampling layer while the classifier contains 3 fully-connected layers with dropout. We used a ResNet-50^41^ as the SAGE model encoder initialized using weights from ImageNet^42^ pretraining (“IMAGENET1K_V1”) and made available through the torchvision model library. An embedding vector of size 256 was used for training the decoder and classifier modules. All models were trained using two 16B Nvidia A4000 GPUs on the DigitalOcean cloud computing service with a virtual 16-core Ubuntu 22.04 machine and 96.6GB of RAM. We trained an ensemble of five SAGE models using the same train split (90%) from the HAM1000 dataset, ensuring no overlap of unique lesions between training and test sets. We completed training in two stages with a batch size of 64 and early stopping with a patience value of 5. An initial warmup stage trained the encoder and classifier using the combined center loss of the latent space embedding and weighted cross-entropy loss of classification until center loss no longer improved (maximum of 50 epochs). The second stage trained the encoder, decoder and classifier for a maximum of 50 epochs and used weighted cross-entropy loss as well as the decoder’s mean-squared error (MSE) of reconstruction. We utilized a learning rate of 1×10^-4^ and a weight decay of 1×10^-5^ with the AdamW^43^ optimizer.

For benchmark analysis, we initialized an ensemble of five ResNet-50 models with ImageNet weights (“IMAGENET1K_V1”) and modified the final fully-connected layer to include dropout. We trained ResNets for a maximum of 50 epochs on the same HAM10000 training split as SAGE with early stopping, weighted cross-entropy loss, a learning rate of 1×10^-4^ and a weight decay of 1×10^-5^ with an AdamW optimizer.

### t-SNE Visualization and Plotting

We generated the latent space embeddings of size 256 for images in all datasets using the trained SAGE models. For visualization purposes only, these embeddings were mapped to t-stochastic neighbor embedding (t-SNE) space using the scikit-learn^44^ (v1.4.2) manifold library, labeled according to the dataset of origin and plotted as scatterplots using matplotlib^45^ (v3.9.0). To explore cluster identities, we plotted HAM10000 train and test images together and colored scatter points according to their diagnostic category. All plots were created using a combination of matplotlib and seaborn^46^ (v0.13.2), and figures were assembled using BioRender.

### Uncertainty Quantification Scoring

Before evaluation, we preprocessed all images with the same steps taken for the training data and used the trained SAGE models to generate three output measures as previously described and detailed in Fig. 2.^33^ For our benchmark analysis, we include the first output measure (average kNN distance) as a standalone measure of uncertainty.

Let *x* = (*x*_1_, *x*_2_, *x*_3_) represent the observed output values for a given image, corresponding to the three SAGE model measures:

1. *x*_1_: L1 latent embedding distance to the *k* = 20 nearest training neighbors
2. *x*_2_: Softmax classifier confidence (argmax)
3. *x*_3_: Reconstruction error

Let *X*_*i*_ be the random variable denoting the distribution of the model’s output for measure *i* across the training data and let *F*_*i*_(*x*) = *P*(*X*_*i*_ ≤ *x*_*i*_) denote the cumulative distribution function (CDF) of *X*_*i*_.

We define the exceedance probability for measure *i* as:

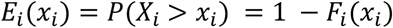

The SAGE score for an image is then computed as the geometric mean of the three exceedance probabilities:

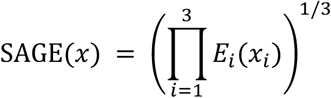

We use the trained ResNet-50 ensemble to generate three additional measures of uncertainty for comparison. First, we evaluate all models on an input image, *x*, and average the softmax predictive distributions:

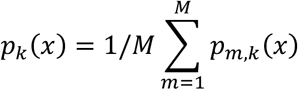

Where *p*_*k*_(*x*) is the ensemble-averaged probability, *p*_*m*,*k*_(*x*) is the softmax probability for class *k* from model *m*. We use the simple baseline of the maximum softmax probability (MSP) from the averaged distributions as the first measure of uncertainty from the ResNet ensemble.

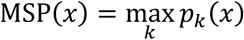

We next calculate mutual information (MI) as a measure of model disagreement:

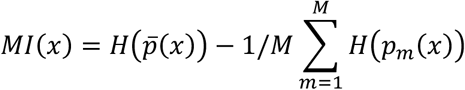

Where *H*(̅*p*(*x*)) is the Shannon entropy of the ensemble-average probability outputs and *H*(*p*_*m*_(*x*)) is the predictive entropy of model *m*. Entropy is defined as:

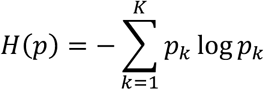

Finally, we evaluate the first trained ResNet model using stochastic dropout (*p* = 0.2) on the fully-connected layer during repeated forward passes (*n* = 5). We then average the resulting softmax predictive distributions and calculate normalized entropy as follows:

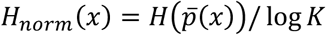

### Image Annotation and Parallel Coordinates Plot

We manually-annotated 12 independent image features (e.g. lesion contrast to skin, presence of non-skin background) that could impact quality and reliability of malignancy prediction as detailed in Extended Table 3 for HIBA, UFES and DDI datasets. Images were reviewed by a single consistent observer (W.M.S.) with clinician oversight (E.B. and R.F.T.). Biasing information such as diagnosis, FST and malignancy status were removed prior to annotation. All annotations were appended to the metadata accompanying each dataset. After annotation was completed, we normalized the observed levels of 5 image features (lesion contrast, camera flash, hair level, presence of ruler and non-skin background) and plotted a line for each test image with the color denoted by the image’s calculated SAGE score. We plotted images in order from low to high-scoring examples and added random jitter of 1.25% to improve visibility of line overlays.

### Comparing SAGE Scores Across Skin Tones for High-Quality Images

Using the manually annotated image features, we removed all images from the HIBA, DDI and UFES datasets with camera flash, measuring devices, high hair density (hair level ≥ 2) and any non-skin background (background ≥ 1) leaving 2,959 high-quality images. We merged FST levels into ‘Light’ (FST I-II), ‘Medium’ (FST III-IV) and ‘Dark’ (FST V-VI) groups and compared the pairwise distributions of SAGE score values using a one-sided Wilcoxon rank-sum test using the scipy^47^ package (v.1.13.0) with the ‘less’ alternative option. We calculated Cliff’s Delta to measure the effect size of distribution non-overlap using a custom python function (see Code & Data Availability).

### OOD Detection Benchmarking

We evaluated HAM10000 test, HIBA, UFES, DDI, MILK10K and Caltech-101 images on trained SAGE and ResNet-50 ensembles and performed UQ as described above resulting in five measures per image: average Maximum Softmax Probability (MSP), ensemble Mutual Information (MI), normalized entropy (ENTROPY) from stochastic dropout, kNN distance to train embeddings (KNN) using the trained SAGE latent space and the average SAGE score. For MSP, KNN and SAGE we take the inverse of score values so all UQ metrics fall in the range [0 - 1] where 0 is most confident and 1 is most uncertain. We mixed in-distribution (HAM10000 test) images with all images matching five severity levels of OOD from most to least severe: 1) semantic shifts where images include object categories but no skin lesions, 2) clinical images with classes not present in the HAM10000 dataset, 3) clinical images with classes shared by HAM10000, 4) dermoscopic images with classes not present in HAM10000 and 5) dermoscopic images with classes shared by HAM10000 but captured in different clinical settings. For each grouping and UQ method we calculate the false positive rate at 95% training recall (FPR95) using the scitkit-learn ‘roc_curve’ function and determine area-under-the-ROC-curve (AUROC) with scikit-learn’s ‘roc_auc_score’ function, with the goal of separating ID from OOD image examples. For both FPR95 and AUROC we report 95% confidence intervals calculated from repeat bootstrapped sampling (*n* = 300).

### Malignancy Prediction

We used a binary (i.e. malignant v. benign) deep-learning predictor from Daneshjou et al.^26^ based on the Inception v3 architecture and pre-trained on HAM10000 images. We implemented this model in python according to instructions on the project’s GitHub repository, as this was the only algorithm from the paper with a publicly-available training dataset (https://github.com/DDI-Dataset/DDI-Code). Preprocessing steps used before malignancy prediction were the same as for training and evaluating our SAGE model. Only basal cell carcinoma, squamous cell carcinoma and melanoma classes were considered malignant for HAM10000 images. The HIBA, UFES, DDI and MILK10K datasets came pre-annotated with histologically confirmed malignancies which were used as ground truth labels. All images with an output greater than or equal to 0.733 were classified as malignant, with those falling below this threshold classified as benign lesions for all datasets. This output threshold was determined as the value that maximized F1 score on the HAM10000 test split. We calculated intrinsic malignancy prediction confidence as a function of the distance to the decision boundary, *b*, using the following equation where *x* is the classifier’s output value for a given prediction and *k* is the scaling variable:

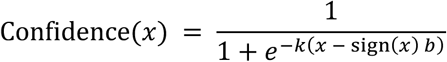

with

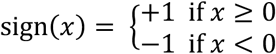

### Risk-Coverage Analysis

After UQ scoring and malignancy prediction was performed for all images, we randomly-sampled images from the five previously-defined ID:OOD groups at a fixed 2:1 ratio with replacement. For each UQ metric, we sorted the sampled dataset by UQ values from low to high and calculated risk (1 - accuracy) across coverage levels (i.e. uncertainty quantiles). This process was repeated for each dataset and UQ metric for 1,000 bootstrapped runs and the median risk-coverage curve and 95% confidence intervals were reported. The resulting curves were plotted using matplotlib and area-under-the-curve was calculated using numpy’s ‘trapz’ function. We repeated this analysis using only intrinsic UQ from the malignancy predictor and compared it directly to SAGE risk-coverage across all ID:OOD mixtures.

### SAGE Thresholding and Receiver Operating Characteristic Curves

Using the HAM10000 training dataset SAGE score distribution, we determined the score value yielding 90% training recall and use it as a binary threshold above which predicted OOD images are filtered. For each skin tone grouping (‘Light’, ‘Medium’, ‘Dark’) and image modality (‘Dermoscopic’, ‘Clinical’), we calculated area-under-the-receiver operating characteristic (AUROC) curves before and after thresholding to assess changes to malignancy model generalization and plotted our results using scikit-learn (‘RocCurveDisplay’) and matplotlib.

## Results

We obtained five publicly-available skin lesion imaging datasets (HAM10000^34^, HIBA^35^, UFES^37^, DDI^26^ and MILK10K^38^) each with distinct characteristics and metadata (Extended Table 1; See Methods). HAM10000, the most widely used dataset for benchmarking, contains only one image size (600×450 pixels) whereas images from other datasets vary in resolution and dimensions (range: 147×147 – 4128×3176 pixels). HAM10000 features lesions with eight diagnostic classes, five considered largely benign (actinic keratosis, benign keratosis, dermatofibroma, melanocytic nevi, and vascular skin lesion) and three considered malignant (basal cell carcinoma, squamous cell carcinoma and melanoma). (Extended Fig. 1) Notably, nearly half of all lesions in the DDI dataset (45.12%) represented 36 other diagnoses not present in the HAM10000 dataset whereas this proportion is small (1.97%) for MILK10K. (Extended Table 2) Both dermatoscope and clinical smartphone imaging modalities are included in the HIBA and MILK10K datasets where the ratio of dermoscopic to smartphone images is 3.67:1 and 1:1, respectively.

Using HAM10000 as a primary reference dataset, we trained five independent SAGE models to compress skin lesion images into 256-dimensional latent space vectors, reconstruct the original images and predict diagnostic class from the compressed embeddings. (Fig. 1) We found that classifier modules performed relatively poorly on HAM10000 holdout test images (balanced accuracy = 0.581) in contrast to decoder modules which were able to reconstruct lesion size, shape and pigmentation color without higher resolution details like hair (Extended Fig. 2). Additionally, the SAGE encoder latent space could distinguish variation within and between different image sets (e.g. HAM10000, training and testing distributions were highly similar whereas HIBA, UFES, DDI and MILK10K occupied areas of training data paucity) (Extended Fig. 3).

**Figure 1.**
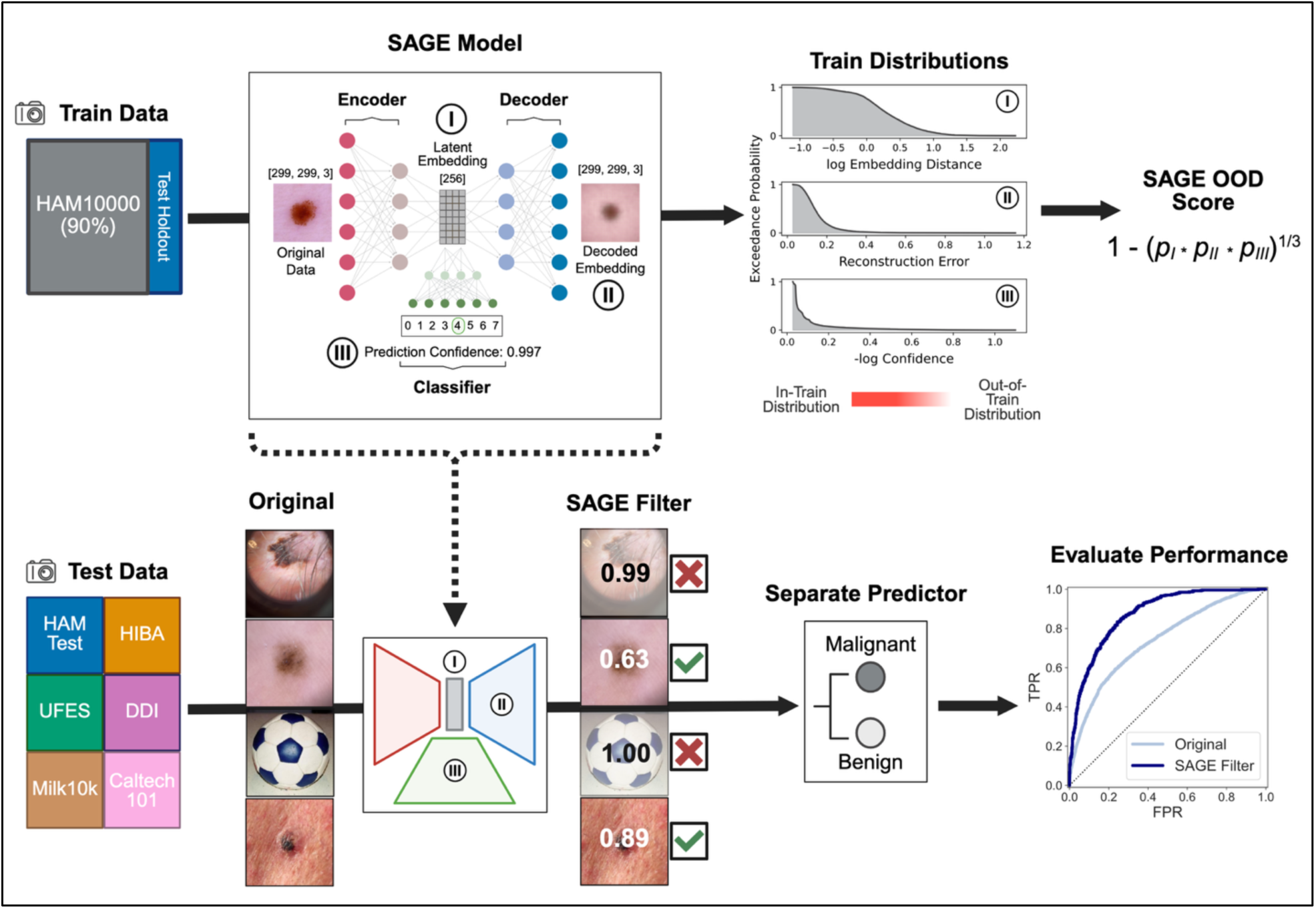
Overview of SAGE model training and deployment for improving performance of a separate malignancy predictor. In the top-left corner, the HAM10000 dataset is split into training and test sets with the training set used to fit the SAGE model’s encoder, decoder and classifier components. The location of the three SAGE components are shown within the model’s greater architecture with each component assigned a label (I-III). To the right, the output values for each component is shown as a labeled distribution corresponding to exceedance probabilities of the training data (Methods). Values move from more likely (“In-Train Distribution”) to less likely (“Out-of-Train Distribution”) to be encountered in the training data as points move from left to right along the x-axis. The SAGE OOD score is shown as the inverse of the geometric mean of the three exceedance probabilities. In the bottom-left corner, 4 images from the test datasets are shown before (‘Original’) and after (‘SAGE Filter’) scoring with the trained SAGE model. Boxes with checkmarks show images with SAGE scores below a chosen threshold, with a red ‘X’ denoting where scores above this amount. A separate, pre-trained model is used to predict malignancy of test images and performance is evaluated via an example Receiver Operating Characteristic (ROC) curve. The original and SAGE score-filtered images demonstrate improvement after high-scoring images are removed from the pool of predicted examples as assessed by area-under the ROC curve.

To quantify conformity between the training and test images, we calculated each image’s SAGE score – a multi-criterion metric for assessing similarity based on the reference distributions for the model’s latent embedding distance, classifier confidence and reconstruction error (Methods) – and averaged results across trained SAGE models. The distributions of SAGE scores for HAM10000 testing and non-HAM10000 skin lesion datasets (i.e. HIBA, UFES, DDI, MILK10K) were significantly higher than the train score distribution, with the HAM10000 test images showing the lowest median score (median = 0.63) and DDI the highest score among dermatology datasets (median = 0.96). (Fig. 2A) These dataset-level differences were largely consistent across diagnoses. (Fig. 2B) However, we observed that images associated with diagnoses missing from the HAM10000 dataset had a higher average SAGE score than those corresponding to diagnoses present in the training data (mean = 0.90 vs. mean = 0.75). When we removed images of test lesions with diagnoses not present in the HAM10000 dataset from DDI and MILK10K, we expected that differences in SAGE score might be clearly delineated by metadata such as imaging modality and generalized skin tone. In fact, we found a surprising degree of overlap in these distributions (Dermoscopic, IQR: [0.73 – 0.93]; Smartphone, IQR: [0.82 – 0.94]; Light skin, IQR: [0.81 – 0.96]; Medium skin, IQR: [0.80 – 0.93]; Dark skin, IQR: [0.83 – 0.96]) pointing to the possibility of other distinguishing image attributes that are unidentifiable from the metadata or lesion type alone. (Fig. 2C-D)

**Figure 2.**
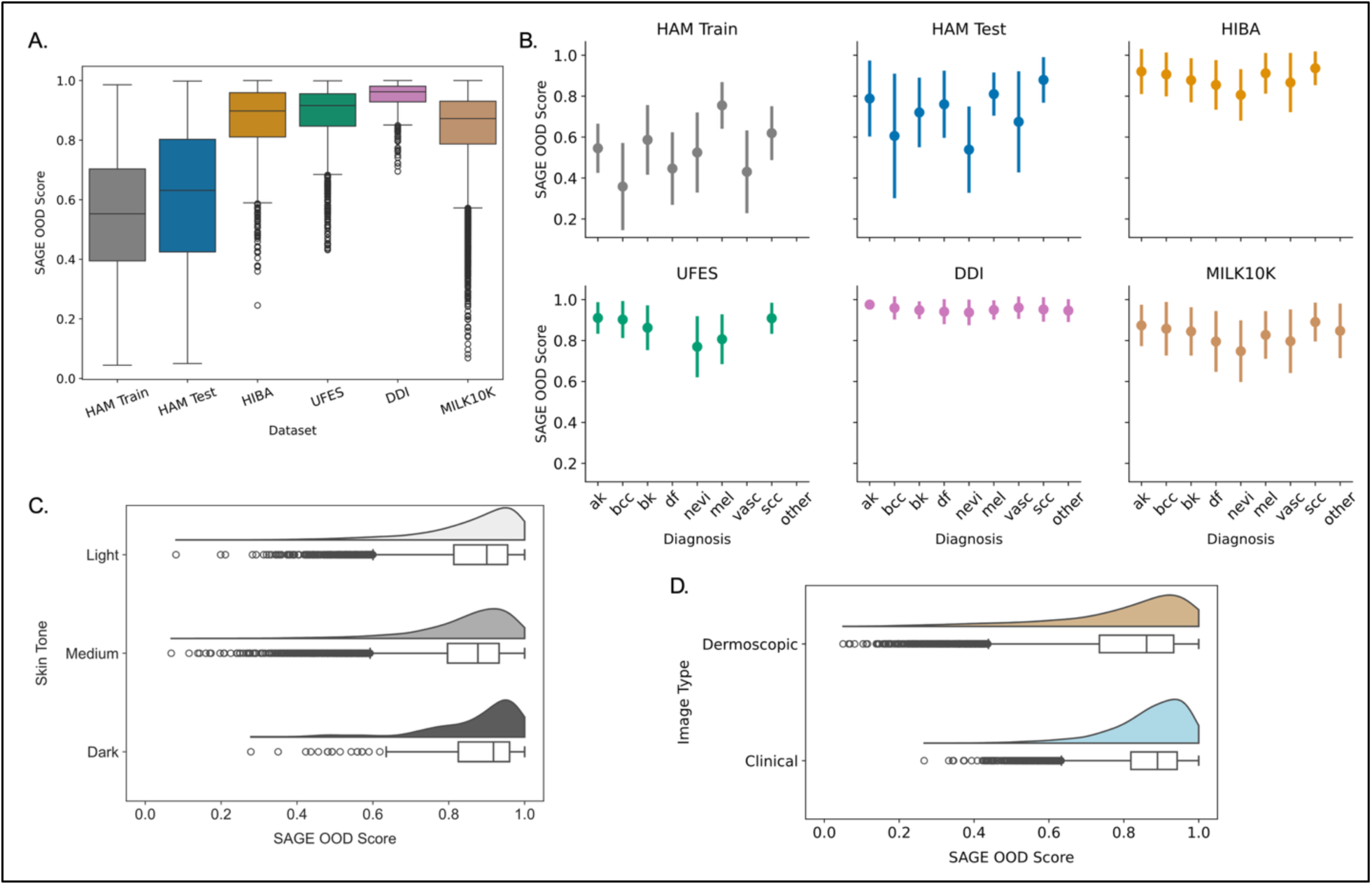
SAGE scores reveal differences between and within dataset categories. A) Distributions of SAGE scores for skin lesion datasets calculated using models fit to the HAM10000 training split are represented as a uniquely-colored boxplot, with the central line denoting the median score values. Top and bottom of each box corresponds to the 25th and 75th percentile of values and whiskers extend to 1.5× the interquartile range (IQR). B) Average SAGE score shown for each of the eight HAM10000 diagnostic categories and split by dataset. Points use the same dataset-specific coloring as in A and categories missing from datasets are omitted (e.g. dermatofibroma not present in UFES data). Images with a diagnostic category not present in HAM10000 data are grouped together as an ‘other’ category. Lines above and below points represent standard deviations of the SAGE score distribution. C) Images from HIBA, UFES, DDI and MILK10K with harmonized skin-tone metadata are split according to three level groupings (Light, Medium, Dark), with SAGE score distributions for each skin-tone group represented as a boxplot and a smoothed kernel density estimate. D) Test images from HAM10000, HIBA and DDI are split according to dermoscopic or clinical smartphone imaging technology, with SAGE score distributions shown as separate boxplots and kernel density estimate. (ak –actinic keratosis, bcc – basal cell carcinoma, df – dermatofibroma, nevi – melanocytic nevi, mel – melanoma, vasc – vascular skin lesion, scc – squamous cell carcinoma)

We note that HIBA, UFES and especially the DDI dataset are laden with high SAGE scoring images with many out-of-distribution features not present in the HAM10000 data, such as measuring devices (e.g. rulers), dense hair, skin coverings and other markings. (Fig. 3C, D, E) Even within the HAM10000 and MILK10K datasets we identified uncommon edge cases with high SAGE scores such as lesions cropped by the image frame or bubbles resulting from a lapsed contact between the dermoscope lens and skin. (Fig. 3A, B, F) To establish how the presence and severity of out-of-distribution features affect SAGE score, we manually-annotated 12 independent features for all HIBA, UFES and DDI images (*n* = 4,527). (Extended Fig. 4, Extended Table 3) Some features had clear positive associations with SAGE OOD scoring such as the presence of a ruler or use of camera flash which showed 7.21% and 5.79% increases compared to the overall mean score for annotated images. Similarly, the presence of non-skin background had a positive effect on SAGE OOD score even at low (7.36%) and medium (10.48%) levels. Linking multiple features together with a SAGE score overlay gives a concise description of what constitutes a typical “high-quality” image, such as having no or low amount of hair, high contrast between the lesion and skin and little to no non-skin background. (Fig. 4A) We frequently observed additive effects when detrimental image features were stacked such as the 11.52% increase to SAGE OOD score when both medium to dense hair and medium to high non-skin background are co-present (mean = 0.99) vs. a 5.13% increase for hair only (mean = 0.93) and a 10.57% increase for majority non-skin background alone (mean = 0.98). (Fig. 4B) We also noted that the presence of some image features did not affect patients with different skin phototypes equally. For instance, inclusion of a ruler had a 6.11% increase to average SAGE score in patients with lighter skin phototypes whereas this increased by 9.77% for patients with dark skin, possibly resulting from the higher contrast between the illuminated measuring devices and darker skin. (Fig. 4C) Intriguingly, while a statistically significant difference between SAGE score distributions for the lightest and darkest skin tones remained after removing low-quality images (one-sided Wilcoxon rank sum test, *p*=0.045), the effect size of non-overlap dropped by over one half from −0.434 to −0.206 (Cliff’s Delta), illustrating how extraneous image features were possibly a larger detriment to image quality than marked changes to underlying phenotypes such as skin tone. (Extended Fig. 5)

**Figure 3.**
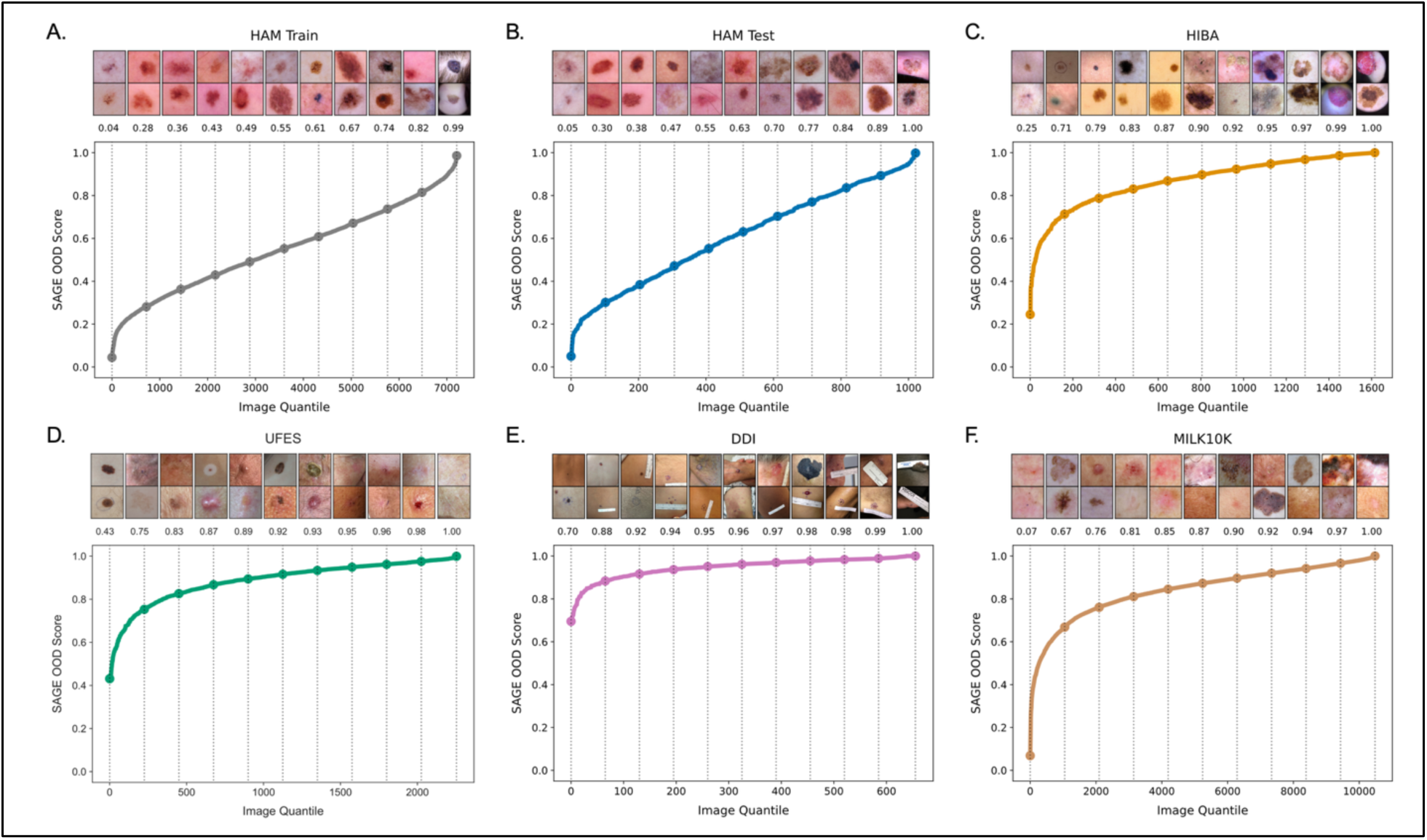
Quantile plots show imaging artifacts associated with high SAGE OOD scores. Images are sorted by SAGE score (y-axis) and plotted by image quantile (x-axis) for A) HAM10000 Train (*n* = 7,207), B) HAM10000 Test (*n* = 1,022), C) HIBA (*n* = 1,616), D) UFES (*n* = 2,255), E) DDI (*n* = 656) and F) MILK10K (*n* = 10,480) datasets. For each quantile plot, a scatter point marks decile intervals and a vertical dotted line leads to the two nearest SAGE score values and the corresponding lesion images after resizing and center crop transforms have been applied.

**Figure 4.**
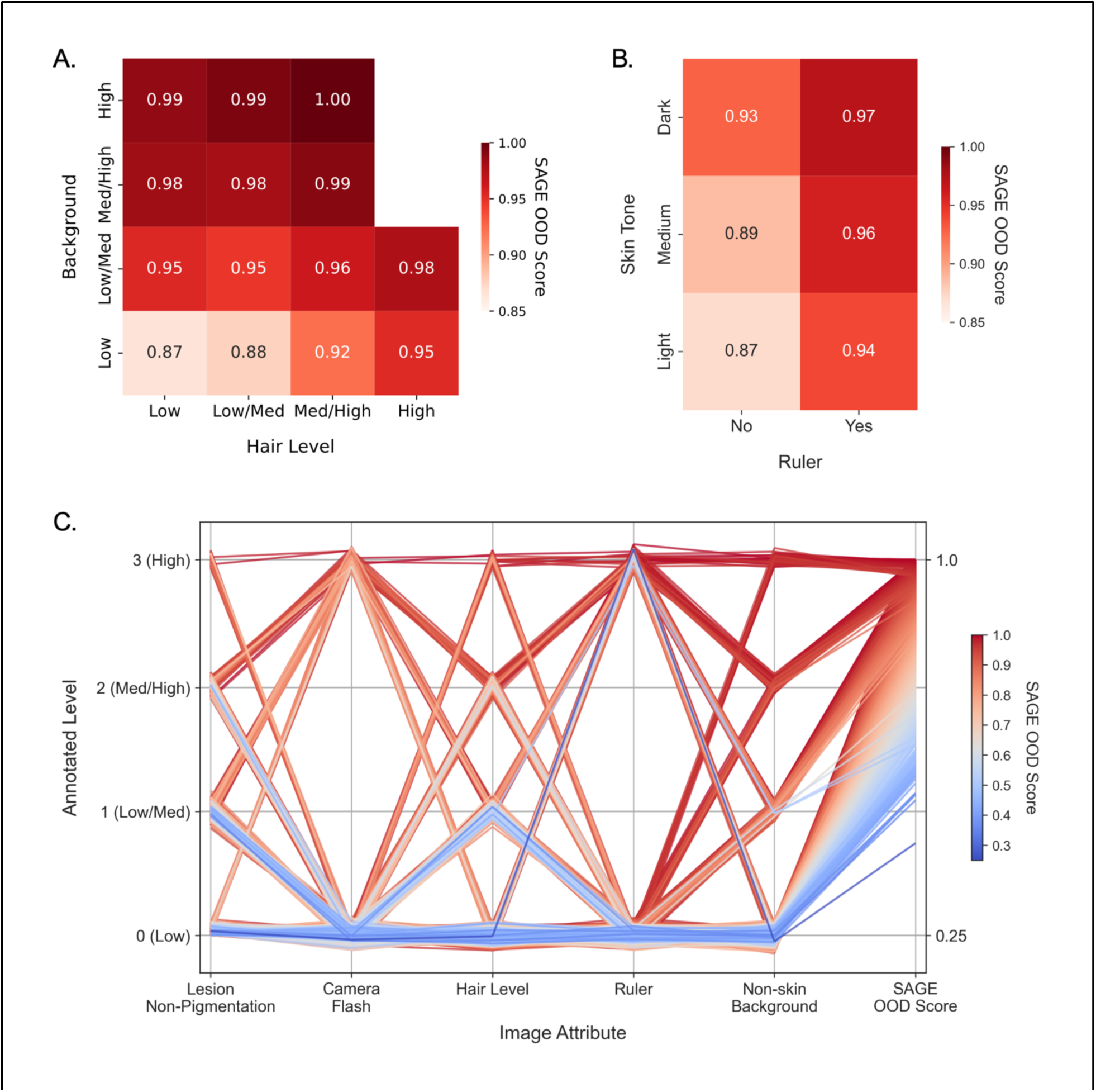
Increasing severity of manually-annotated image features correlates with increased SAGE scores. A) Heatmap of SAGE score by manually-annotated hair density (x-axis) and non-skin background (y-axis) feature severity. Squares are colored based on the average SAGE score value shown in the center of each square, with color intensity determined by the colorbar gradient to the right of the heatmap. Each row corresponds to one severity level of non-skin background and displays the change in average SAGE score as hair density increases from left to right. B) Heatmap of SAGE score by skin type and measuring device levels. Test images are grouped into three categories based on generalized skin tone in three levels (Light, Medium, Dark) and displayed along the y-axis. The x-axis displays the binary presence of a measuring device (ruler) in the lesion image, with heatmap squares displaying the average SAGE score for images at the intersection of the two categories. C) Five manually-annotated attributes for HIBA, UFES and DDI images (*n* = 4,527) are shown along the x-axis with the scaled level plotted on the left y-axis, with the right y-axis showing SAGE score from low (top) to high (bottom). Each image’s combination of features is represented as a continuous line and is assigned a color based on its SAGE score value, with intensity determined by the colorbar gradient to the plot’s right.

We next evaluated the ability of SAGE to detect OOD image examples from a mixed ID and OOD evaluation dataset, a critically important precursor to medical image model generalization. We created mixed datasets with HAM10000 test images and examples from five severity levels of distribution shift from strong (most OOD) to weak (most ID). Note that we introduce semantically unrelated images from the Caltech-101 dataset to assess far-OOD detection. To compare SAGE performance with well-established deep learning methods for UQ, we trained an ensemble of ResNet-50 models (*M* = 5) and estimated classification uncertainty using average maximum softmax probability (MSP), mutual information (MI) from ensemble confidence and normalized entropy (ENTROPY) from Monte-Carlo dropout repeats of the first ResNet’s forward pass (*n* = 5). Additionally, we included the average latent distance to training *k*-Nearest Neighbors (KNN) score derived from our trained SAGE encoders. Our benchmark analysis revealed that SAGE is the top or second top performer for OOD detection on all mixed datasets, and excelled at detection of non-semantic shifts (far-OOD; Area-Under-the-ROC-Curve (AUROC) = 1.00 [0.99 – 1.00], False Positive Rate at 95% Train Recall (FPR95) = 0.01 [0.01 – 0.02], 95% CI) and combined modality and class shifts (OOD; AUROC = 0.92 [0.91 – 0.93], FPR95 = 0.31 [0.27 – 0.36]). (Table 1) As anticipated, detection performance dropped as mixed datasets became more similar to HAM10000 training images, with best FPR95 reaching 63% and AUROC dropping to 0.81 for the expected ID group. We also noted the SAGE ensemble performance is superior to any individual ensemble member for OOD detection. (Extended Fig. 6)

**Table 1.** Benchmark analysis of OOD detection task across five levels of difficulty. The table is split into two sections, with the top showing the False Positive Rate at 95% Recall (FPR95) and the bottom showing the Area-Under-the-ROC (AUROC) for detecting out-of-distribution (OOD) images within a mixed dataset. Rows correspond to uncertainty quantification methods with columns denoting mixed in-distribution (ID) and OOD datasets over increasing difficult to detect shifts. The overall metric is reported using the full mixed dataset with the 95% confidence intervals shown in brackets for each statistic.

|  | FPR95 ↓ |  |  |  |  |
| --- | --- | --- | --- | --- | --- |
|  | Far-OOD | OOD (Class + Modality) | Near-OOD (Modality) | Near-OOD (Class) | Expected ID |
| MSP | 0.54 [0.51, 0.58] | 0.58 [0.54, 0.62] | 0.61 [0.58, 0.64] | 0.63 [0.42, 0.71] | 0.70 [0.67, 0.73] |
| MI | 0.55 [0.52, 0.58] | 0.61 [0.58, 0.67] | 0.64 [0.61, 0.67] | 0.63 [0.43, 0.69] | 0.71 [0.68, 0.74] |
| ENTROPY | 0.99 [0.98, 1.00] | 0.61 [0.51, 0.72] | 0.62 [0.58, 0.66] | 0.77 [0.63, 0.85] | 0.78 [0.76, 0.81] |
| KNN | 0.16 [0.14, 0.18] | **0.29 [0.23, 0.33]** | **0.46 [0.42, 0.50]** | 0.58 [0.38, 0.83] | 0.66 [0.63, 0.69] |
| **SAGE (ours)** | **0.01 [0.01, 0.02]** | 0.31 [0.27, 0.36] | **0.46 [0.43, 0.50]** | **0.56 [0.44, 0.84]** | **0.63 [0.59, 0.66]** |
|  | AUROC ↑ |  |  |  |  |
|  | Far-OOD | OOD (Class + Modality) | Near-OOD (Modality) | Near-OOD (Class) | Expected ID |
| MSP | 0.79 [0.77, 0.80] | 0.68 [0.65, 0.70] | 0.75 [0.73, 0.76] | 0.80 [0.76, 0.84] | 0.72 [0.71, 0.74] |
| MI | 0.79 [0.77, 0.81] | 0.61 [0.58, 0.64] | 0.66 [0.64, 0.68] | 0.78 [0.74, 0.81] | 0.70 [0.68, 0.71] |
| ENTROPY | 0.48 [0.46, 0.50] | 0.81 [0.78, 0.83] | 0.82 [0.80, 0.83] | 0.72 [0.66, 0.77] | 0.72 [0.70, 0.74] |
| KNN | 0.93 [0.92, 0.94] | 0.89 [0.87, 0.91] | **0.85 [0.84, 0.87]** | **0.84 [0.80, 0.88]** | 0.80 [0.79, 0.82] |
| **SAGE (ours)** | **1.00 [0.99, 1.00]** | **0.92 [0.91, 0.93]** | **0.85 [0.84, 0.86]** | 0.81 [0.77, 0.84] | **0.81 [0.79, 0.82]** |

Beyond OOD detection, UQ can be used to selectively filter images with high risk of model failure in medical settings. Although SAGE contains a classifier module, it is unclear to what degree this and other UQ methods from classification-aligned tasks can improve selective prediction when information on downstream model confidence is restricted. To that end, we performed a risk-coverage analysis with all benchmarked methods and OOD severity mixtures held at a constant 2:1 ID to OOD ratio to enable comparison across groups. We sorted images using OOD scoring functions from low to high before passing to a separate binary malignancy prediction model^26^, finding that most UQ methods produce risk-coverage curves with overlapping 95% confidence bands and share similar Area-Under-the-Risk-Coverage (AURC) values across all dataset mixtures. (Fig. 5) We note that SAGE yields the joint lowest AURC values for mixed datasets with class and modality shift (OOD; AURC = 0.06), only modality shift (Near-OOD, Modality; AURC = 0.10) and Expected ID (AURC = 0.10) mixtures. Intriguingly, intrinsic UQ derived from the malignancy prediction model itself was a weaker predictor of model failure risk than SAGE when encountering mixed datasets with modality shift or those with expected ID images from HIBA and MILK10K datasets. (Extended Fig. 7) We also tested the effect of SAGE score thresholding on model bias against patients with dark skin, finding that a threshold value at 90% Train recall (τ = 0.816) improved performance of dark skin images (Original: AUROC = 0.68; Threshold: AUROC = 0.78) to a level greater than that of light skin (Original: AUROC = 0.75; Threshold: AUROC = 0.77) and medium skin (Original: AUROC = 0.70; Threshold: AUROC = 0.73) images but with heavily reduced coverage (16.42%). (Extended Fig. 8)

**Figure 5.**
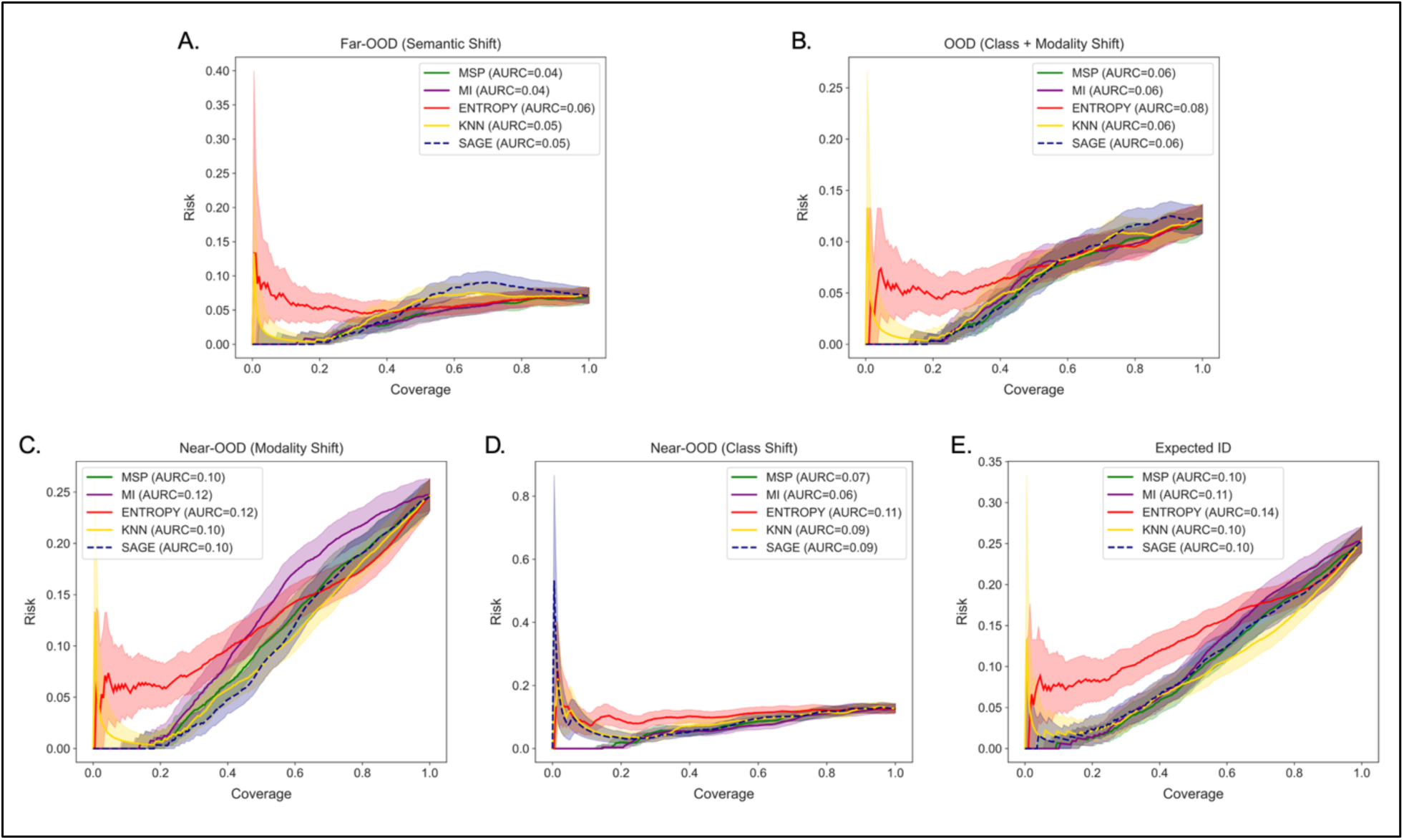
Risk-coverage curves for benchmark UQ methods paired with a downstream malignancy prediction model. Plots display risk (1 - accuracy) of malignancy prediction failure on the y-axis and coverage (proportion of total images being evaluated) on the x-axis for each mixed ID:OOD dataset. Mixed datasets were randomly-sampled with replacement for 1,000 bootstrapped runs, with lines representing the median risk-coverage curve of each benchmarked UQ method and the shaded area showing the 95% confidence interval. Area-under-the-risk-coverage curve (AURC) is calculated and displayed next to each UQ method in plot legends. (MSP – Maximum Softmax Probability, MI – Deep Ensemble Mutual Information, ENTROPY – MC Dropout Entropy, KNN – k-Nearest Neighbors)

Finally, we tested images of previously-unseen malignant lesion types such as T-cell lymphomas, kaposi sarcoma and metastatic carcinoma on the pre-trained malignancy prediction model and found that they had poor zero-shot performance. (Extended Fig. 9A) Surprisingly, these lesions tended to have lower, not higher, intrinsic uncertainty scores than malignant classes featured in the training data making selective prediction on these classes difficult. (Extended Fig. 9B) Using SAGE, we set an OOD score threshold (τ) with 90% recall on the HAM10000 training data (τ = 0.816) and filtered all malignancies falling above this threshold level. We found an enrichment of false negative (FN) images above the OOD score cutoff for all new malignant classes except “Malignant – Other” from MILK10K. (Extended Fig. 9C) Overall, previously-unseen malignancies showed a high proportional capture of FN examples when thresholding on SAGE score before applying the downstream prediction model (macro-average FN capture = 0.81).

## Discussion

In this study we demonstrate the successful application of our novel UQ method to enhance OOD image detection across a spectrum of severity levels and reduce risk of subsequent skin cancer malignancy prediction in a global cohort. Our work uses the most popular benchmarking dataset of dermoscopic lesion images, HAM10000, as a reference to quantify OOD test images and identify detrimental image attributes without selection bias or requiring a classification label. To our knowledge, this is the first study to apply deep ensembles to dermatology imaging datasets with the goal of UQ for dataset comparison. By incorporating five datasets from six countries in our analysis, we also address the critical need to analyze generalization gaps when applying skin cancer prediction models across global populations. Our method can pair with any dermatology ML task so long as the training data is known, which will improve integration of robust UQ into more trustworthy automated pipelines. Additionally, we demonstrate that rich image embeddings derived from supervised and unsupervised signals can be utilized to reduce risk of model failure better than an intrinsic uncertainty score from the downstream model itself in some instances. We envision SAGE as a powerful extension to or replacement for the concept of model cards, where users can view detailed information about a model’s training data.^48^ In our case, we not only quantify the differences between training data and prospective test data but also encourage users to interactively probe examples to see where and how they might differ from distributions of training image features.

However, this analysis has several limitations. Despite sourcing images from five independent datasets, the vast majority of images pertain to patients with lighter skin phototypes (FST I-IV, MILK10K automated skin-tone 4 and 5) and lack significant representation from Asian and African populations. We also note that the FST scale itself has been shown to vary based on environmental conditions.^49,50^ The data in this study consists of a relatively small cohort of 7,207 training and 16,029 test skin lesion images; inclusion of larger imaging datasets or pre-training on dermatological images could strengthen encoder fine-tuning and yield more relevant embeddings instead of initializing on default ImageNet weights from PyTorch. Additionally, the training and testing sets may contain different lesions from the same patient. We are also unable to assess the influence of identifiable markings such as scars or tattoos, as these were explicitly removed by the authors of each benchmarking dataset for patient de-identification purposes. We did not manually-annotate features for HAM10000 or MILK10K images due to large dataset size, and the incidence of these features in the SAGE training data was not analyzed, although automated annotation methods are available that could expedite this process in future work.^51^ Furthermore, other image features that have been shown to affect ML model performance on skin imaging datasets such as color balance (e.g. blue or red pixel intensity shifts) were not annotated or assessed. Additionally, we did not study the effects of anatomical location on SAGE scoring which could provide additional guidance to users of the downstream malignancy prediction model.

While our model architecture and latent vector size of 256 dimensions was selected to yield embeddings of sufficient quality, we did not explore larger latent spaces (e.g. >500 dimensions) or other model architectures which might facilitate improved image reconstructions and better classification performance. As hardware support for training large ML models continues to decrease in cost, our work could expand to encompass larger foundational encoders and more complex latent space embeddings. Future work will also seek to pair SAGE with more difficult image segmentation, skin cancer diagnosis and lesion monitoring tasks across patient groups and will test a larger array of downstream models to develop an interval of task improvement facilitated by SAGE. Finally, while this work was explicitly focused on skin cancer detection using dermoscopy and clinical photography, SAGE could be broadly applicable for a wide array of clinical and non-clinical use cases.

## Data Availability

All data are publicly available using the download links provided in the Methods section. Project code and trained models are freely available via our GitHub repository link.

https://github.com/pdxgx/sage

## Code & Data Availability

All datasets used in our analysis are publicly available and download links are included in Methods. Code for SAGE model training and retrieving SAGE scores is freely available on our GitHub repository (https://github.com/pdxgx/sage).

## Author Contributions

W.M.S planned the analysis, wrote the code and wrote the main manuscript text. R.S. reviewed and edited the manuscript. E.B. reviewed and edited the manuscript and provided insights related to dermatology practice. R.F.T. planned the analysis, edited the manuscript and provided funding.

## Extended Figures

**Extended Table 1.** Overview of dermatology imaging datasets. Table rows correspond to key metadata categories for each dataset including country of origin, lesion and patient demographic statistics. Each unique dataset is assigned its own column. Missing category information is denoted as “unknown”.

|  | HAM10000 | HIBA | UFES | DDI | MILK10K |
| --- | --- | --- | --- | --- | --- |
| Origin | Austria, Australia | Argentina | Brazil | United States | Austria, Australia, North Macedonia, Turkey, United States |
| Total Images | 10,015 | 1,616 | 2,298 | 656 | 10,480 |
| Unique Patients | *unknown* | 623 | 1,373 | 570 | *unknown* |
| Unique Lesions | 7,470 | 1,246 | 1,641 | *unknown* | 5,240 |
| Imaging Technology | Dermoscope | Dermoscope, Smartphone | Smartphone | Smartphone | Dermoscope, Smartphone |
| Male / Female % | 54.30 / 45.70 | 46.90 / 53.10 | 49.60 / 50.40 | *unknown* | 60.15 / 39.85 |
| Light Skin % (FST I-II) | *unknown* | 80.00 | 41.60 | 36.70 | 27.65 |
| Medium Skin % (FST III-IV) | *unknown* | 20.00 | 55.20 | 31.70 | 70.23 |
| Dark Skin % (FST V-VI) | *unknown* | 0.00 | 3.20 | 31.60 | 2.12 |

**Extended Figure 1.**
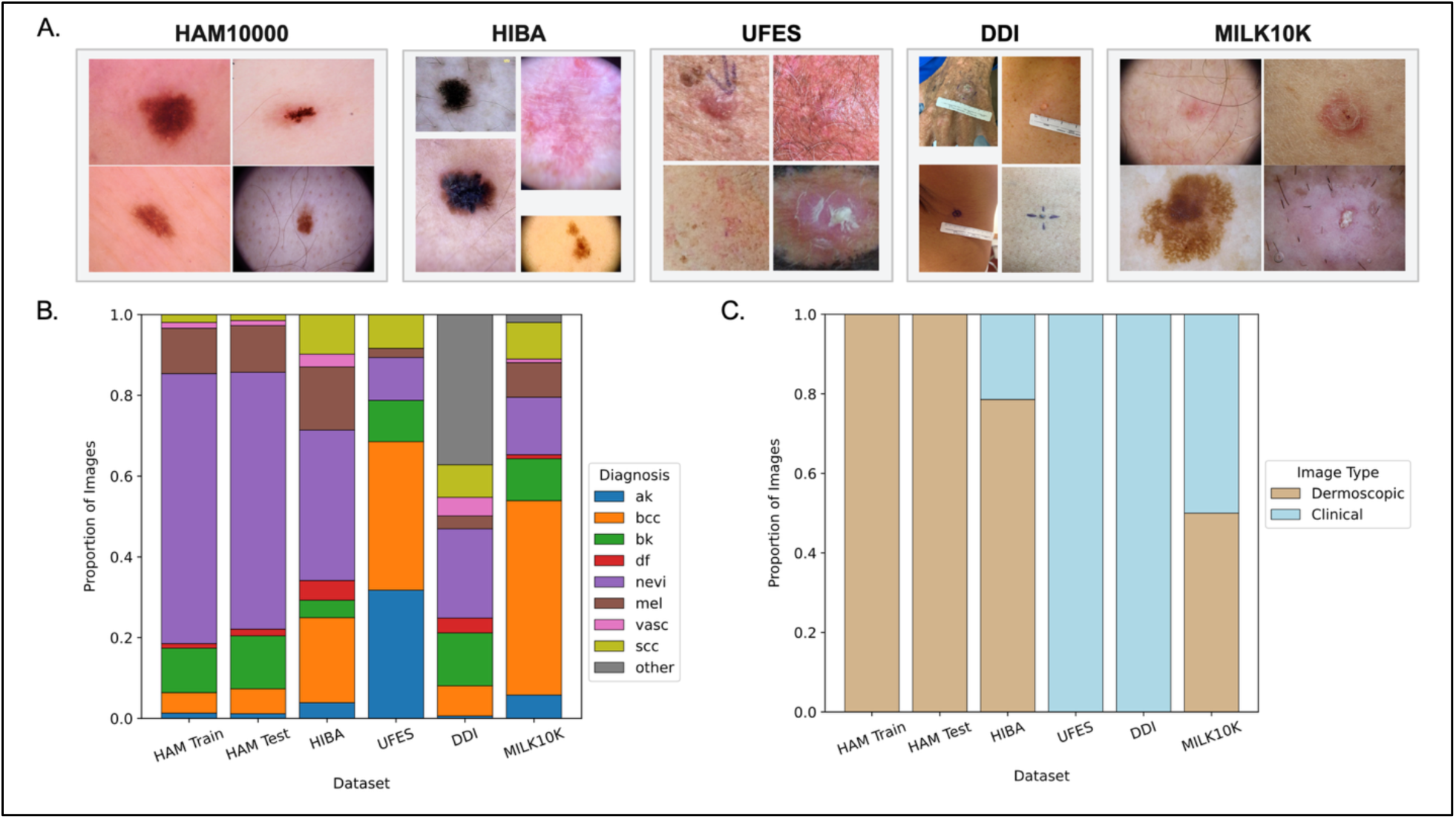
Visualization of differences in dermatology dataset metadata. A) Each unique dataset is represented as a rectangle with a random selection of 4 images showcasing dataset-wide variations in image proportions, technology, image angle. Dermoscopic images from HAM10000, HIBA and MILK10K show head-on perspectives with lesions centered in the image frame whereas UFES and DDI exhibit a greater variety of lighting and angle changes with the introduction of clinical smartphone imaging. B) Stacked barplots of the proportion of datasets corresponding to diagnostic categories. Each dataset is shown on the x-axis with a stacked barplot where the proportion of all diagnoses sums to 1. The eight diagnoses from the HAM10000 train and test data are represented by colored bars while additional diagnoses not present in the HAM10000 dataset are shown as a gray ‘other’ category. C) Stacked barplots showing the proportion of imaging modality present in each dataset. X-axis labels correspond to unique datasets and the total proportion of images sums to 1. (ak –actinic keratosis, bcc – basal cell carcinoma, df – dermatofibroma, nevi – melanocytic nevi, mel – melanoma, vasc – vascular skin lesion, scc – squamous cell carcinoma)

**Extended Table 2.**
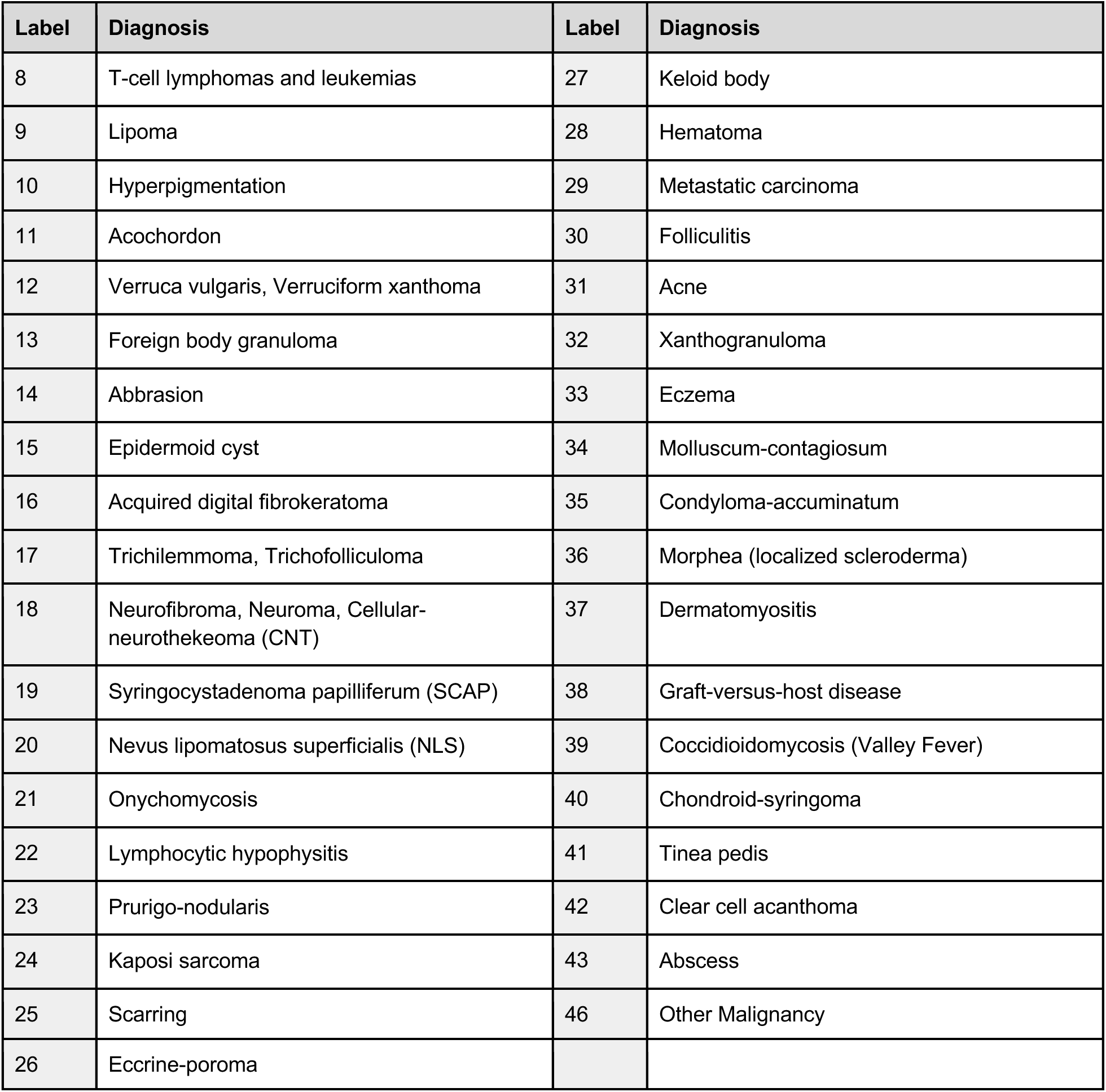
Lesion diagnoses missing from HAM10000 dataset. Table of diagnoses present in the HIBA, UFES and DDI datasets that are not present in the HAM10000 training and test data. Labels start from the end point of the eight HAM10000 classes [0-7]. Classes 8-43 are only present in DDI with class 46 only found in MILK10K. Where applicable, similar diagnoses are grouped into the same label category such as wart-like lesions (12) and benign nerve cell tumors (18).

**Extended Figure 2.**
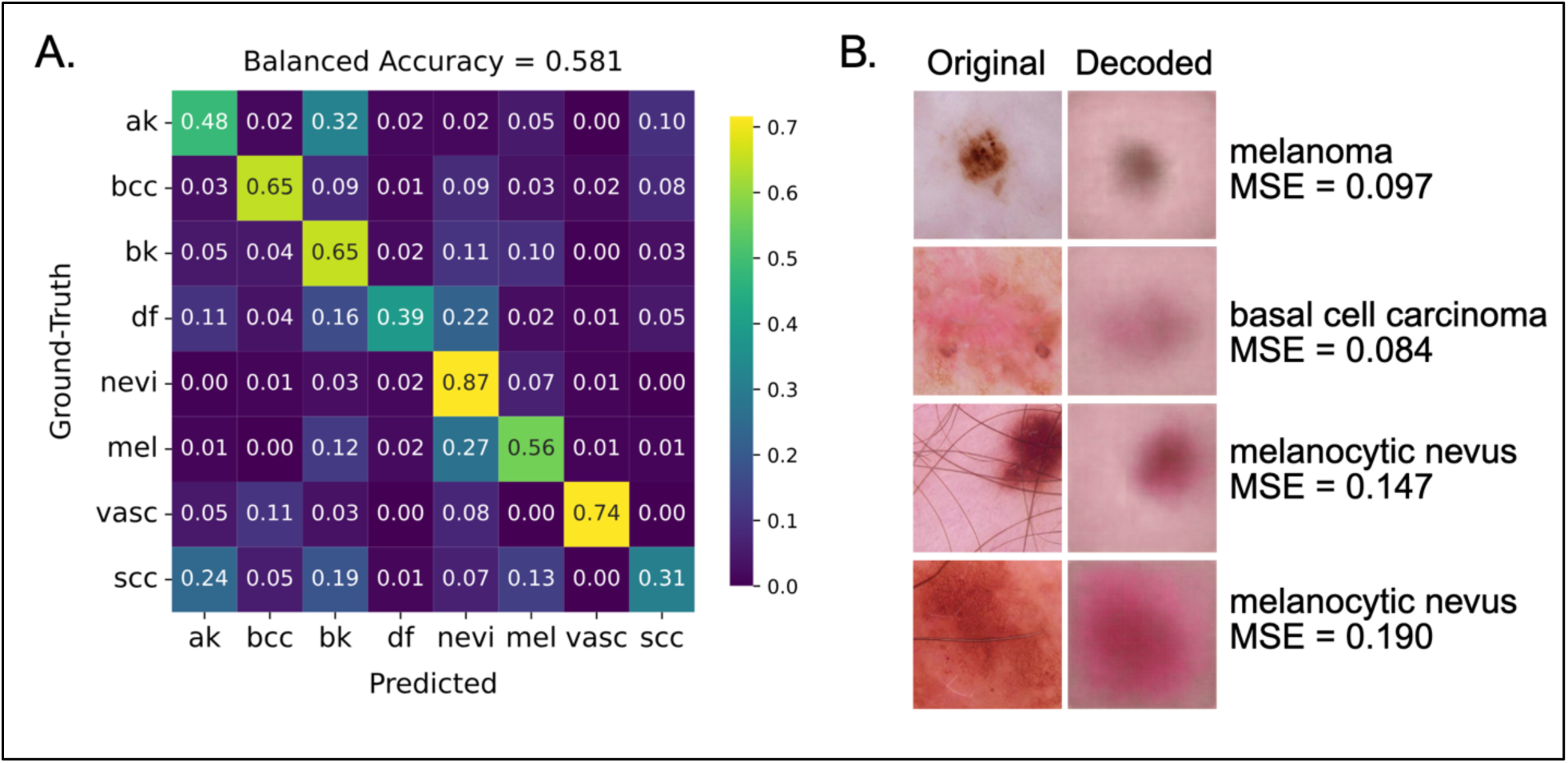
Performance of SAGE model classifier and decoder on HAM10000 test images. A) Confusion matrix of trained SAGE model classifier predictions (x-axis) versus ground truth diagnostic labels (y-axis). Each row displays the predicted classes for a ground-truth label, with squares showing the number of images for each predicted class. Squares are colored according to the accuracy values in the colorbar to the right of the confusion matrix and the overall balanced accuracy is displayed above the plot. B) Original images from the HAM10000 test set are displayed in the left-hand column with the result of the embedding and decoding process using the trained SAGE model displayed in the right-hand column. For each row, the diagnostic label is displayed as well as the mean-squared error (MSE) of the decoded image.

**Extended Figure 3.**
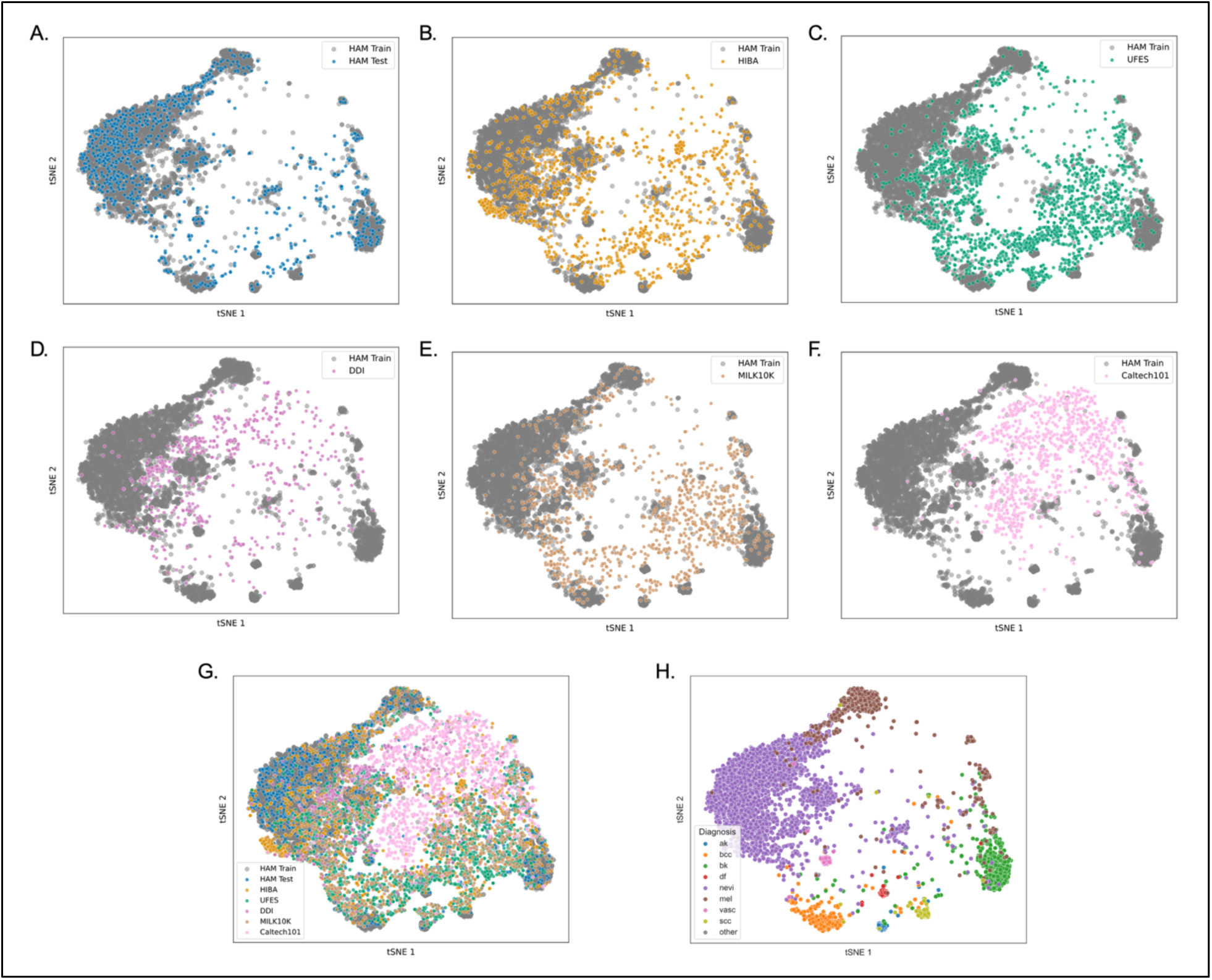
Visualization of 256-dimensional embedding space with t-SNE. A-F) Images for HAM10000, HIBA, UFES, DDI, MILK10K and Caltech-101 test datasets are compressed using the trained SAGE model encoder, with the resulting embeddings projected into t-SNE space. HAM10000 train image embeddings are plotted in gray to illustrate the reference embedding space after dimensionality reduction with t-SNE. G) Overlay of all dataset embeddings in t-SNE space. H) Images from all datasets are projected into t-SNE space and colored according to ground-truth label, with clusters arising based on diagnosis such as melanocytic nevi (*nevi*) and melanoma (*mel*). Images with a diagnosis not present in the HAM10000 dataset are labeled in gray as an ‘other’ category.

**Extended Table 3.** Description of manually-annotated image features. Rows correspond to 12 independent features used to manually-annotate all HIBA, UFES and DDI images. Each row contains a brief written overview, the variable type and descriptions of up to 4 levels of severity for ordinal variables and 2 levels for binary variables.

| Attribute | Description | Variable | Level 0 | Level 1 | Level 2 | Level 3 |
| --- | --- | --- | --- | --- | --- | --- |
| Size | Lesion size w.r.t. cropped image frame | Ordinal | No discernable lesion | Very small to small size, nearly all skin | Majority of image is skin, lesion visible | Majority of image is lesion, skin visible in background |
| Contrast | Lesion contrast with skin | Ordinal | No contrast | Lesion barely visible, blends with skin | Lesion clearly visible | Lesion starkly visible against skin background of different color |
| Hair Level | Hair density present in image | Ordinal | No hair | Some hair visible | Denser hair (arm, chest, partial scalp) | Very dense hair (scalp) |
| Skew | Off-center position of lesion in image | Ordinal | Lesion centered | Some off-center skew | Lesion on periphery of image or cropped | – |
| Non-skin Background | Background visible in image | Ordinal | No non-skin background | Any non-skin visible in image background | ¼ - ½ of image consists of non-skin background | Over ½ of image is non-skin background (dermoscope border, clinic) |
| Cover | Cloth or bandage covering of skin | Ordinal | No skin covering | Any skin covering visible | ¼ - ½ of skin is covered | Over ½ of skin in image is covered (hospital gown, bandage) |
| Marking | Colored marker on skin denoting lesion location | Binary | No marking | Marking present | – | – |
| Ruler | Measuring device in image | Binary | No ruler | Ruler present (white, black, green) | – | – |
| Flash | Camera flash used | Binary | No flash | Flash used | – | – |
| ENEM | Eyes, nose, ear or mouth visible in image | Binary | Not present | Present | – | – |
| Blurry | Unfocused image | Binary | Image focused | Image unfocused | – | – |
| Multiple | More than one lesion of similar size present | Binary | One lesion | Multiple lesions | – | – |

**Extended Figure 4.**
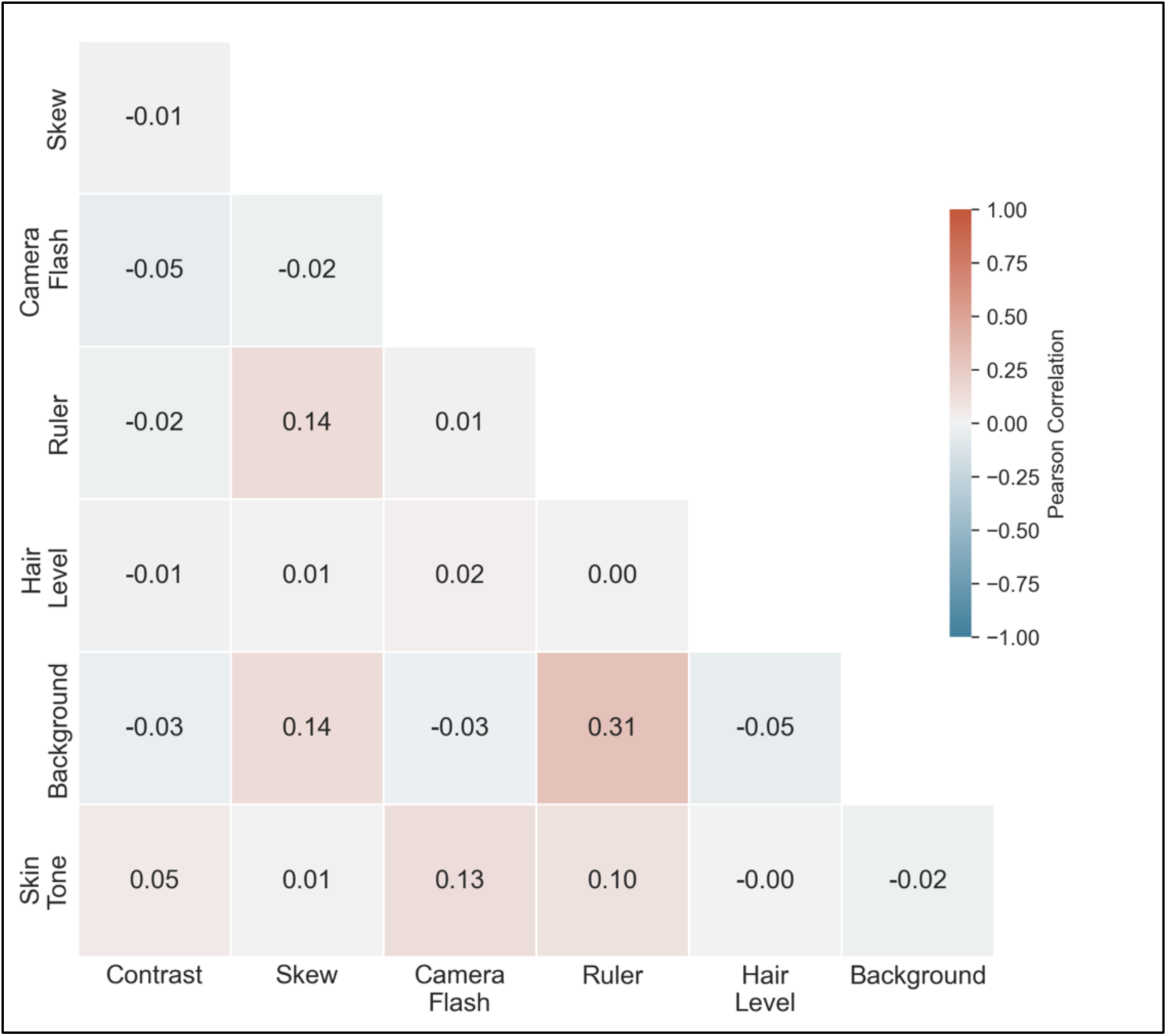
Correlation analysis of manually-annotated image features. A lower triangular matrix displays the Pearson correlation values of 7 unique manually annotated image features for the combined HIBA, UFES and DDI datasets. Each square contains the calculated correlation coefficient and a color based on the gradient shown in the color bar to the right of the matrix. Positive coefficients signify a positive relationship whereas a negative sign shows a negative relationship, with values under 0.3 considered to be a weak correlation.

**Extended Figure 5.**
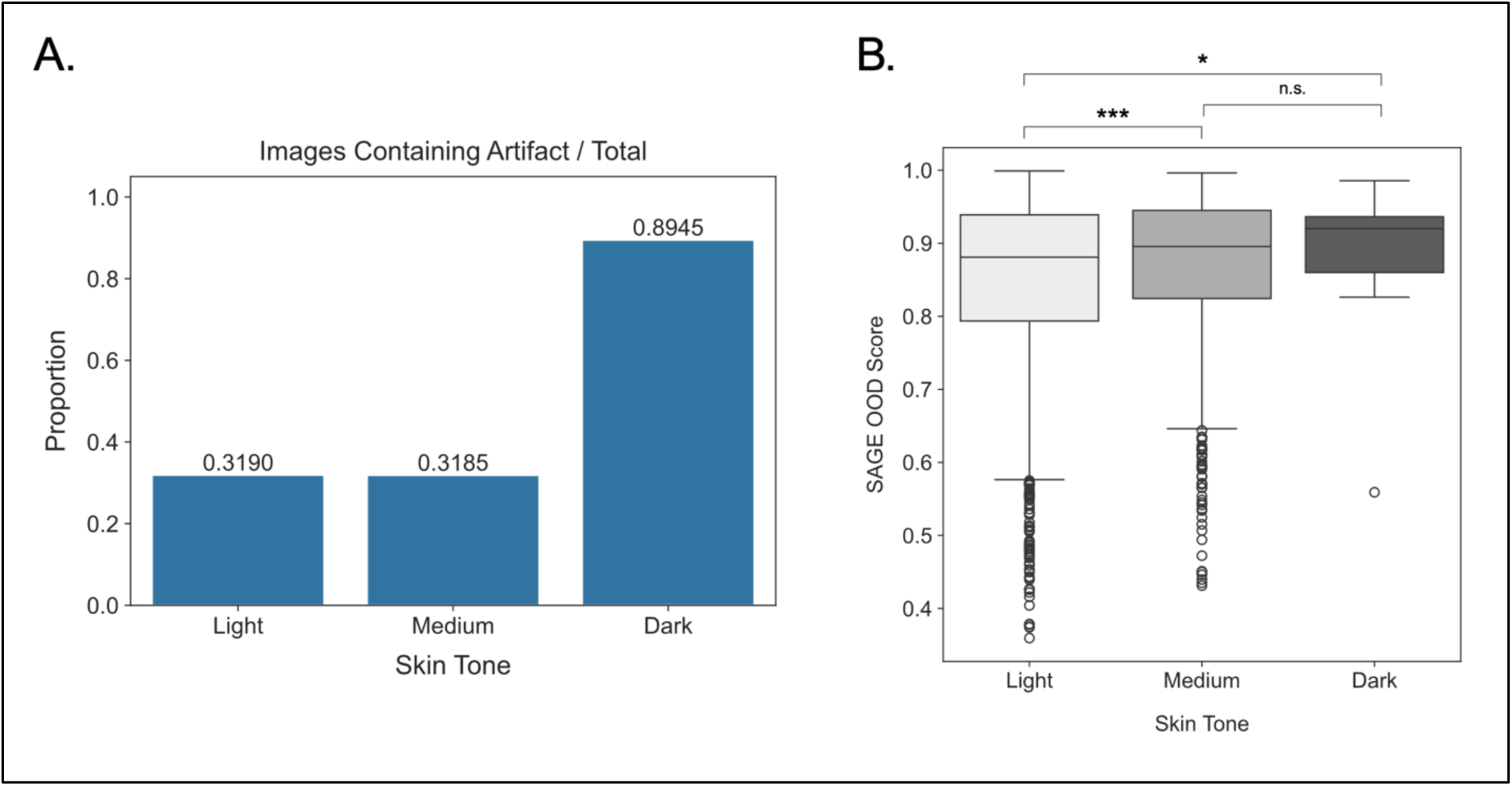
SAGE score distributions for skin tone levels display higher degree of overlap after filtering low-quality images. A) Barplots displaying proportion of skin lesion images containing an extraneous artifact (camera flash, measuring device, medium or high hair density or non-skin background) on the x-axis and skin tone group on the x-axis. Artifact proportion is displayed as decimals above each bar. B) SAGE score distributions for HIBA, UFES and DDI datasets are combined and split by skin tone after filtering examples with manually-annotated artifacts indicative of low image quality (Methods), with distributions shown as boxplots and statistical significance between distributions calculated using a one-sided Wilcoxon rank-sum test (α = 0.05). SAGE score differences are significant between Light and Dark (*p* = 0.045) and Light and Medium skin (*p* = 1.317×10^-5^) and with no significance (n.s.) found between Medium and Dark skin (*p* = 0.174). (*: *p* < 0.05, **: *p* < 0.01 ***: *p* < 0.001)

**Extended Figure 6.**
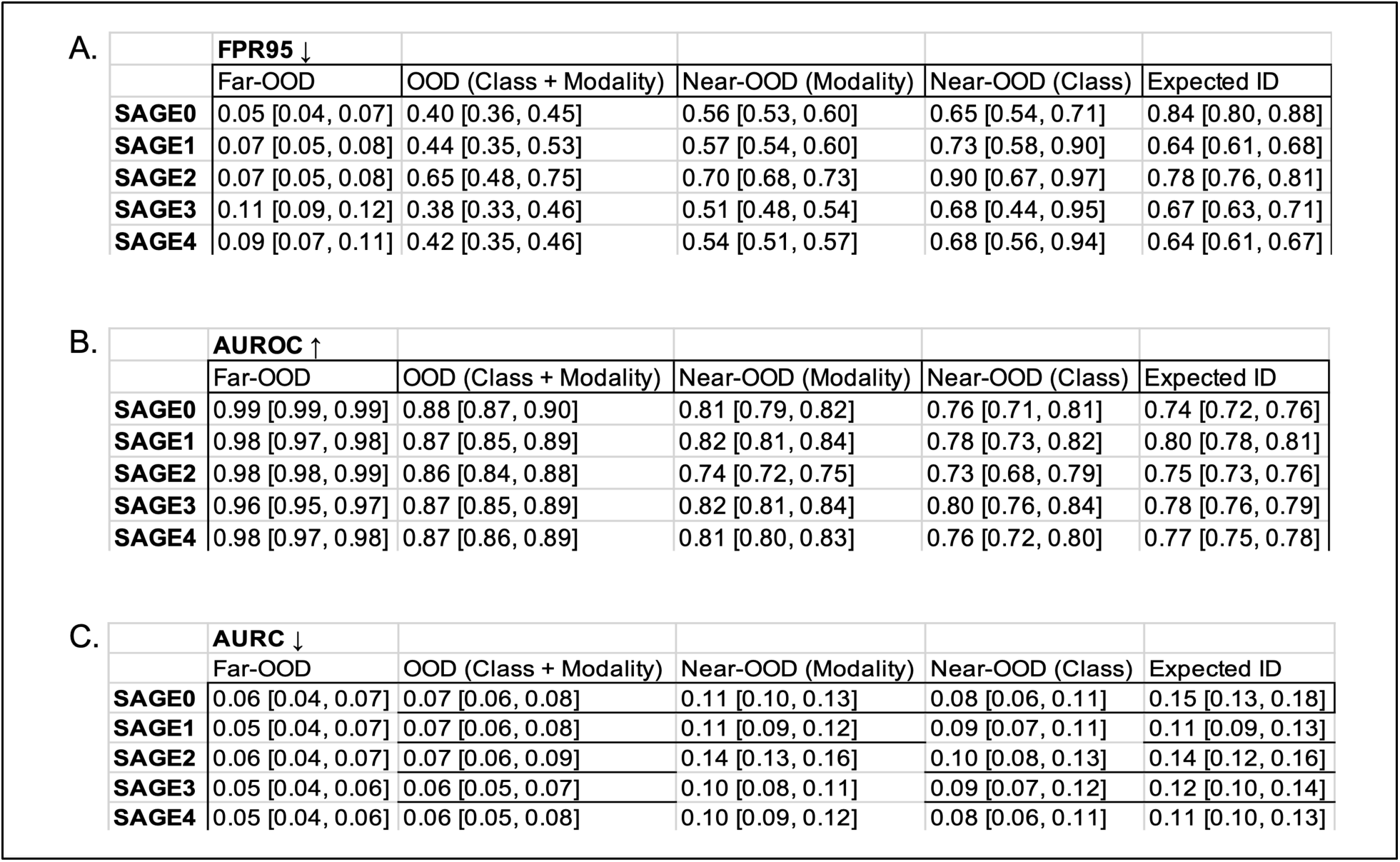
Individual performance of SAGE ensemble members across OOD detection and selective prediction tasks. Each row corresponds to a trained SAGE model and each column denotes a mixed ID:OOD group at increasing difficulty from left to right. Brackets denote 95% confidence intervals determined by bootstrap sampling with replacement. A) FPR95 of detection. B) AUROC of detection. C) AURC from selective prediction task where ID:OOD held at a constant 2:1 ratio.

**Extended Figure 7.**
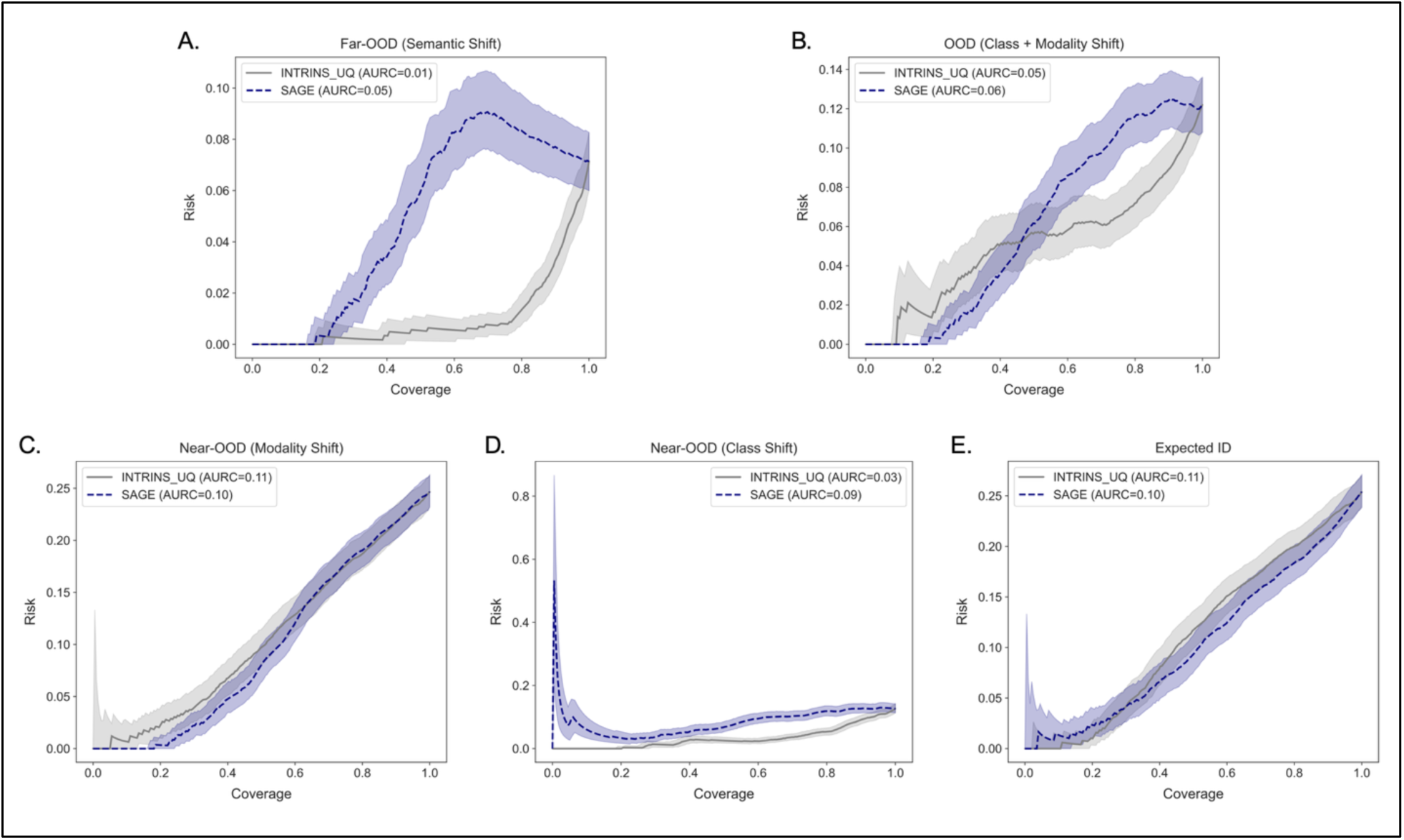
Comparison of risk-coverage curves from intrinsic malignancy prediction model uncertainty versus SAGE OOD scoring. Plots display risk (1 - accuracy) of malignancy prediction on the y-axis and coverage (proportion of total images being evaluated) on the x-axis for each mixed ID:OOD dataset. Mixed datasets were randomly-sampled with replacement for 1,000 bootstrapped runs, with lines representing the median risk-coverage curve of each benchmarked UQ method and the shaded area showing the 95% confidence interval. Area-under-the-risk-coverage curve (AURC) is calculated and displayed next to each UQ method in plot legends.

**Extended Figure 8.**
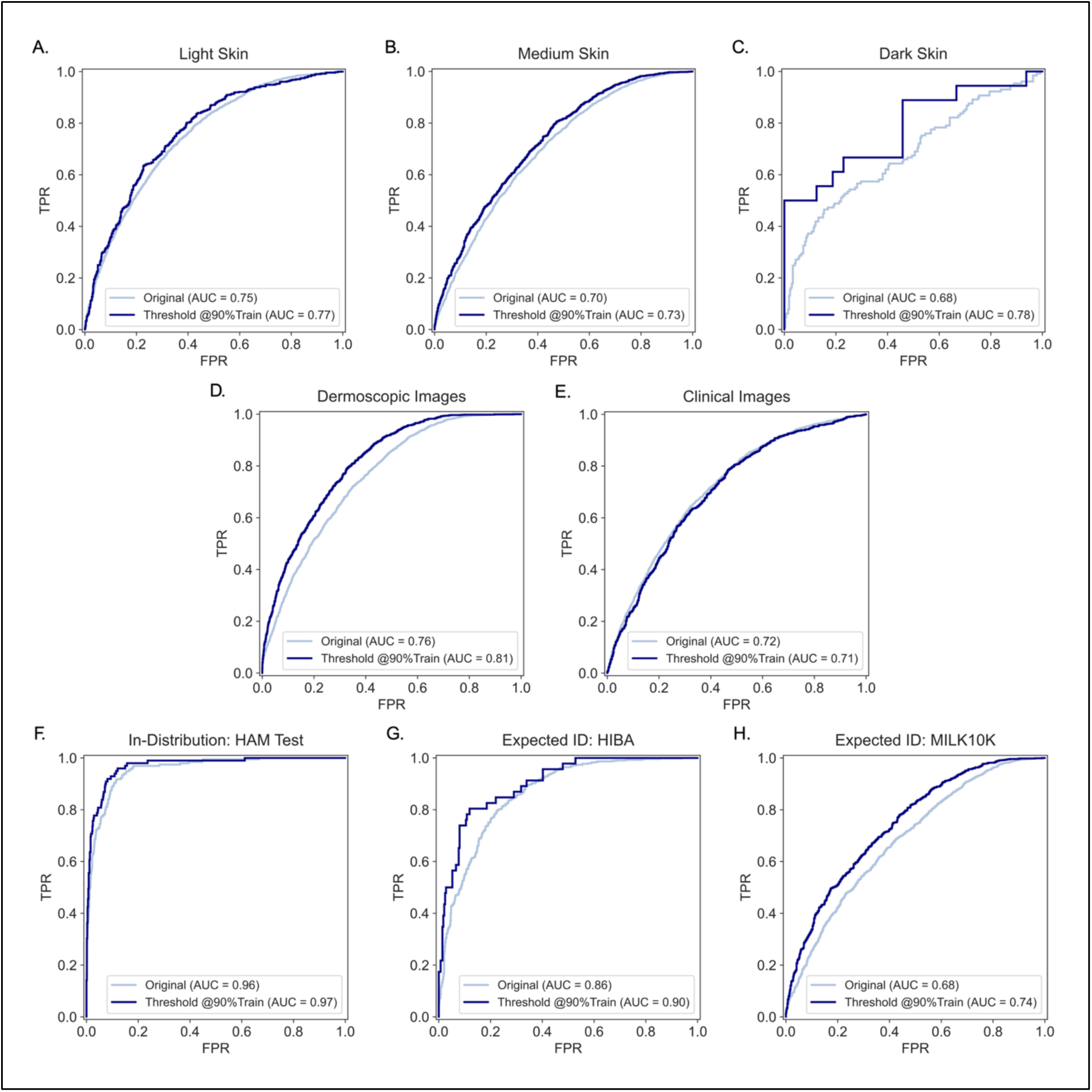
ROC curves show improvement of separate malignancy predictor performance for key metadata categories. A-C) Test images for HIBA, UFES and DDI are split by generalized skin tone and ROC curves are shown after filtering examples below a SAGE score threshold values with 90% Train dataset recall (τ = 0.816). An ROC curve is plotted for the original dataset and for the dataset after SAGE score thresholding for the separate malignancy predictor’s raw output values, with the false positive rate shown on the x-axis and true positive rate displayed on the y-axis. Area-under the ROC curve (AUC) is displayed beside the value in the legend. D-E) Test images for HAM10000, HIBA, UFES, DDI and MILK10K are split by imaging technology and filtered above the same SAGE score threshold, with ROC curves generated using retained images below each threshold. F-H) ROC curves before and after SAGE thresholding for the most ID datasets, with curves for HAM10000 Test, HIBA and MILK10K plotted with dermoscopic images and ID classes.

**Extended Figure 9.**
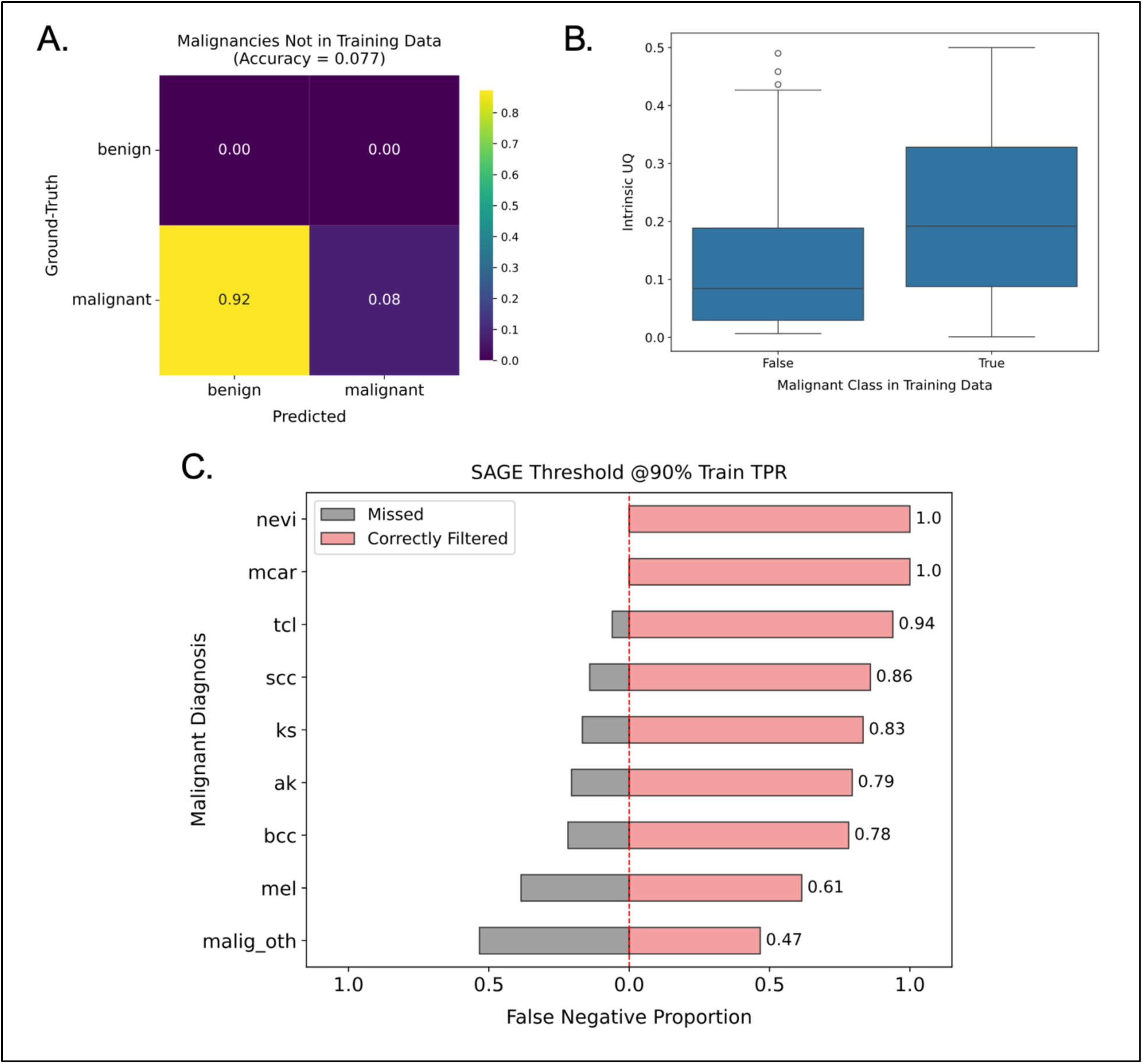
False negative malignancies are selectively filtered with a SAGE score threshold. A) Confusion matrix showing predicted versus ground truth labels for test images of malignant lesion classes not featured in the HAM10000 training data. B) Barplots showing the distribution of intrinsic uncertainty of the downstream prediction model for test malignancies from classes featured versus not featured in the training data. C) False negative (FN) predictions for each disease class from all test skin lesion datasets below the cutoff value of 0.733 are split into two groups: those above and below a SAGE score threshold of 0.816 where the HAM10000 training data shows 90% recall. The x-axis is centered at 0.0 and the red bars right of the center line show the absolute proportion of FN examples falling above the SAGE score threshold for each disease class (“Correctly Filtered”). Bars to the left of the center line show the proportion of FN images that are below the SAGE score threshold in grey (“Missed”). (nevi – melanocytic nevi, mcar – metastatic carcinoma, tcl – T-cell lymphomas, scc – squamous cell carcinoma, ks – kaposi sarcoma, ak – actinic keratosis, bcc – basal cell carcinoma, mel – melanoma, malig_oth – other MILK10K malignancy including collisions)

